# Evidence-to-decision frameworks: a review and analysis to inform decision-making for environmental health interventions

**DOI:** 10.1101/2021.05.04.21256541

**Authors:** Susan L. Norris, Max T. Aung, Nicholas Chartres, Tracey J. Woodruff

**Author notes:** **Corresponding author:** Nicholas Chartres, Program on Reproductive Health and the Environment, Department of Obstetrics, Gynecology & Reproductive Sciences, University of California San Francisco, San Francisco, California, United States. **Declaration of competing financial interests:** Susan L Norris is a member of the GRADE Working Group and has published on GRADE; she is a former employee of the World Health Organization and in that role contributed to the WHO Handbook for Guideline Development (2nd edition 2014) and to the WHO-INTEGRATE framework. Max T. Aung: Nothing to declare Nicholas Chartres: Nothing to declare Tracey J Woodruff: Nothing to declare.

## Abstract

**Background:** Evidence-to-decision (EtD) frameworks provide a structured and transparent approach for groups of experts to use when formulating recommendations or making decisions. While extensively used for clinical and public health recommendations, EtD frameworks are not in widespread use in environmental health.

**Objectives:** This review sought to identify, compare and contrast key EtD frameworks for decisions or recommendations on interventions used in clinical medicine, public health or environmental health. Our goal was to identify best practices and guidance which will be used to inform the development of an EtD framework for formulating recommendations regarding interventions to prevent or mitigate the harmful effects of exposure to substances in the environment.

**Methods:** We identified a convenience sample of EtD frameworks used by a range of organizations. We searched Medline for systematic reviews of EtD frameworks used in clinical medicine, and public or environmental health. In a qualitative manner, we summarized the decision criteria in the selected frameworks and in the reviews.

**Results:** Fourteen key organizations provided 18 EtD frameworks; most frameworks focused on clinical medicine or public health interventions; four focused on environmental health and three on economic considerations. Only one framework was based on an underlying conceptual model, and rarely was a systematic review of potential criteria performed during the frameworks development. GRADE encompasses a set of closely related frameworks for different types of decisions. Harms of interventions were examined in all frameworks and benefits in all but one. Other criteria included certainty of the body of evidence (15 frameworks), resource considerations (15), feasibility (13), equity (12), values (11), acceptability (11), and human rights (2). There was variation in how specific criteria were defined. The five identified systematic reviews reported a similar spectrum of EtD criteria.

**Discussion:** The EtD frameworks examined encompassed similar criteria, with tailoring to specific audience needs. However, there is variation in development processes, terminology, level of detail provided and presentation of the criteria. Existing frameworks are a useful starting point for development of one tailored to decision-making in environmental health.

## Introduction

It is widely recognized that environmental pollution is an important determinant of health and that interventions, in particular policy recommendations, are a key means by which population health can be improved. The formulation of trustworthy and impactful recommendations and policies on environmental health interventions is a complex task, which requires the identification of all relevant data and evidence, their critical appraisal and synthesis and translation into a recommendation or policy. The processes and methods for hazard identification and risk assessment of environmental substances based on systematic reviews of the evidence have advanced considerably in the last decade.^1–4^ These approaches have been largely based on methods developed for clinical medicine,^5^ which have been expanded to encompass public health interventions,^6^ diagnostic test accuracy and impact,^7^ coverage decisions,^8, 9^ and health technology assessments (HTA),^10^ among others.

The final step in the process of formulating evidence-informed recommendations and policies involves the translation of data and evidence on various decision factors into an explicit recommendation or policy. Evidence-to-decision (EtD) frameworks provide a structured and transparent approach for groups of technical experts or policy-makers to accomplish this step.^6, 11^ These frameworks include explicit criteria which the group considers individually and in aggregate focused on the relative benefits and harms, as well as other considerations. Such frameworks can facilitate: i) consideration of all relevant criteria in the decision-making process; ii) examination of the pros and cons of each intervention option; iii) presentation of relevant evidence for each criterion; iv) identification of the reasons for any disagreement within the expert group; v) transparent reporting of the decision-making process; and vi) crafting of the rationale statement for each recommendation. Populated EtD frameworks can also facilitate implementation in the local context, by assisting the end-user in understanding how and why specific recommendations were made, and by providing data and evidence on each decision criterion which may help to facilitate local adoption or inform adaptation.

In environmental health, once hazards are identified and risks assessed, organizations may want to examine mitigating and prevention interventions and make recommendations and policies based on systematic reviews of research evidence and other data and information. An EtD framework suitable for environmental health interventions will facilitate this process. However, such frameworks have not been widely used in this field.

The objectives of this review were to identify, compare and contrast key EtD frameworks for interventions in clinical medicine, public health and environmental health and to summarize the key decision criteria across these frameworks. The identification of these key elements will inform the development of an EtD framework for use in formulating recommendations regarding mitigating and prevention interventions related to exposure to harmful substances in the environment.

## Methods

In order to identify relevant, existing EtD frameworks, we took two approaches: a search of the peer-reviewed literature for systematic reviews of EtD frameworks, and identification of frameworks used by a range of organizations which make decisions or formulate recommendations in clinical medicine, or public or environmental health.

### 1. Scope

The focus of this paper is on the substantive criteria for decision-making with respect to interventions, including both normative criteria (what should be done) and feasibility criteria (what can be done).^12^ Criteria related to the process (versus substance) of decision-making are beyond the scope of this work. While criteria in healthcare decision-making can be either qualitative or quantitative,^10^ this paper focuses on the former, although quantitative criteria (commonly used in HTA) were not excluded.

There are an increasing number of organizations which use systematic and transparent approaches to synthesize evidence in environmental health for hazard identification,^13^ risk assessment,^14^ and for the synthesis of the benefits and harms of interventions.^15^ These organizations were not included in the current analysis because they do not make recommendations on interventions to mitigate the effects of harmful exposures.

### 2. Bibliographic database search and article screening

To identify relevant EtD frameworks, we searched Medline via PubMed for systematic reviews of EtD frameworks, templates or tools published in English and indexed in the 10-year period up to 7 January 2021. The complete search strategy is found in Annex 1. We also solicited the advice of guideline development experts for any additional potential citations. Publications were included if they were systematic reviews of frameworks, tools or templates for formulating decisions or recommendations, or for priority setting related to the use of interventions (including diagnostic tests) in clinical medicine, public health, or environmental health. The included frameworks focused on population-level interventions; decision tools for the provider-patient interaction and patient decision aids were excluded. Frameworks which focused exclusively on economic considerations were also excluded. The setting for both decision-making and implementation of recommendations was not restricted.

Two persons (SLN and MTA) screened the titles and abstracts, and full-text versions were retrieved for studies which potentially fulfilled inclusion criteria. Consensus was achieved between the two reviewers for final inclusion in the review.

### 3. Search for EtD frameworks used by key organizations

in addition to the systematic review described above, we examined a broad range of organizations including clinical and public health guideline development groups in academia and the private sector, healthcare provider professional organizations, governmental agencies and international organizations. Given the purpose of this review, we focused particularly on organizations that work in the field of environmental health. Organizations were identified based on author knowledge of prominent guidelines in public health and clinical medicine, by snowballing (reviewing the origin of or basis for identified frameworks), and by conferring with a broad network of guideline developers internationally based on our experiences and contacts.

We used this pragmatic approach for several reasons. First, because of the dominance of the GRADE (Grading of Recommendations Assessment, Development and Evaluation) EtD framework over the last 15 years,^5, 11, 16^ many organizations either use GRADE or a modification thereof. Thus, a more exhaustive search was unlikely to yield novel or unique frameworks. Second, because the methods used by guideline development organizations are infrequently published in the peer-reviewed literature, an extensive hand-search of the grey literature including organizational web-sites would have been required, which was infeasible.

### 4. Data extraction and synthesis

For each systematic review identified in our search, a single author (SLN) extracted key information describing the focus of the review, time period searched, and the main findings, including an overview of the criteria identified across the included frameworks. We did not extract data on each individual framework included in each review.

For the EtD frameworks used by key organizations, we extracted data on organizational characteristics, the methods used to develop the framework, the funders and any declared interests of the developers of the framework, how evidence should be used to inform each criterion in the framework, quality assessment of individual studies and of the body of evidence, the specific EtD criteria, and the types of conclusions or recommendations formulated. One author extracted these data (SLN) and a second checked them (MTA); disagreements were discussed and consensus reached. Data were extracted into a template in Excel (Microsoft Corporation, Redmond WA, USA)

For the key organizations, the primary decision criteria for each EtD framework were extracted from the main published reports, and specifically from the identified template or list of criteria. For organizations for which there are significant background materials or publications such as for the GRADE framework, the decision criteria that were extracted into the tables and figure include those that are explicitly presented as part of the framework. Other concepts or criteria which are mentioned only in the narrative text accompanying a framework were not extracted unless the text suggested that they were important and were consistently applied in decision-making. Where a clear set of criteria could not be identified (e.g., criteria were presented in narrative text without clear demarcation of the main decision criteria, or when criteria appeared to vary across publications), the authors of the framework were contacted for clarification.

Data were summarized in a narrative, qualitative manner. Given that the organizations and frameworks discussed were a convenience sample, descriptive statistics and statistical comparisons are not meaningful. Quality assessment of the identified EtD frameworks was not performed as there is no standard for such an assessment.

## Results

### 1. Identified systematic reviews of EtD frameworks

The bibliographic database search for systematic reviews of EtD frameworks yielded 399 citations, of which four fulfilled inclusion criteria^7, 10, 17, 18^ (Table 1, Annex 2 and 3). One additional study which fulfilled inclusion criteria was identified by the co-authors.^8^ Each of these reviews included a cohort of EtD frameworks which the authors had systematically identified, with a particular focus: multicriteria decision analysis in HTA,^10^ vaccine adoption into national programs,^17^ diagnostic tests,^7^ decision-making in local low-income settings,^18^ and frameworks for informing health system coverage decisions.^8^ Each review summarized the main EtD criteria identified across their included, individual frameworks, and these main criteria are presented in Table 1. All five reviews included the balance of benefits and harms and consideration of resource use or cost-effectiveness. Only the review by Mustafa and colleagues^7^ included an assessment of certainty of evidence. Considerations of equity, acceptability and feasibility were included only in the reviews by Baltussen et al.^10^ and Burchett et al.^17^ The EtD criteria outlined in these five reviews generally corresponded to the main criteria in the EtD frameworks of the selected key organizations (see Results section 2). One additional publication, excluded from our review because it was not a systematic review, provides a useful list of “value assessment frameworks” for use in HTA for clinical interventions.^19^

**Table 1.** Systematic reviews of evidence-to-decision frameworks: review characteristics and key findings. Decision criteria are the main criteria that the review authors identified across the frameworks which were included in their review. Abbreviations: EtD, evidence-to-decision; GRADE, Grading of Recommendations, Development and Evaluation; HTA, health technology assessment; MCDA, multi-criteria decision analysis; n, number of studies; NR, not reported

|  | Citation | Focus | Years searched; databases | Findings | Decision criteria | Comments |
| --- | --- | --- | --- | --- | --- | --- |
| 1 | Baltussen et al. 2019 (10) | Systematic review of MCDA and related terms; focused on HTA: included only studies that included economic analyses | 1990 to September 2018; Medline only | n=36 studies; categorized these as qualitative (n=1 study), quantitative (n=35), and MCDA with decision rules (n=0); provided list of included studies but no other details | <u>Criteria for MCDA:</u><br>1. Effectiveness<br>2. Severity of disease<br>3. Disease of the poor<br>4. Cost-effectiveness | Search strategy focused only on MCDA and studies that included economic analyses; no data provided on the 36 individual studies |
| 2 | Burchett et al. 2012 (17) | Systematic review of the literature on national decision-making about adoption of new vaccines into national immunization programs | Through March 2010; Medline and multiple other databases; multiple languages | n=21 unique frameworks | <u>Nine broad categories of criteria:</u><br>1. Importance of the health problem (eg disease burden)<br>2. Effectiveness and safety of the vaccine<br>3. Programmatic considerations<br>4. Acceptability<br>5. Accessibility, equity and ethics<br>6. Financial/economic issues<br>7. Impact of vaccination<br>8. Consideration of alternative interventions<br>9. Decision-making process | Provides list of domains and sub-domains (Table 2); for vaccine context |
| 3 | Morgan et al. 2018 (8) | Systematic review of EtD frameworks focusing on decision-making about whether or not to pay for a new healthcare intervention (e.g., test, treatment, or procedure) | 2013-2015; multiple databases, English only | n=25 frameworks, each with a set of decision criteria | Variable across the 25 frameworks.<br><u>Developed a new framework, built on GRADE EtD, including:</u><br>1. Burden of disease<br>2. Benefits and harms<br>3. Values and preferences<br>4. Resource use<br>5. Equity<br>6. Acceptability<br>7. Feasibility<br>Modifications included adding limitations of alternative technologies considerations in use (expanding benefits and harms) and broadening acceptability and feasibility constructs to include political and health system factors. | Started with GRADE for clinical interventions, modified it for coverage/payer decision-making; did not examine the EtD criteria for all 25 identified frameworks. |
| 4 | Mustafa et al. 2017 (7) | Systematic review to identify tools for assessing the quality of evidence and the strength of recommendations related to diagnostic strategies and tests in health care | 1996 to June 2012; Medline, Embase | Identified 29 tools and 14 modifications | <u>Over all tools examined, domains to assess strength of recommendations:</u><br>1. Quality of evidence<br>2. Patients and populations beliefs<br>3. Cost and resources<br>4. Balance of benefits and harms/burden | Focus on diagnostic tests only; Table 3 includes categories for EtD criteria (with various sub-domains or synonyms) |
| 5 | Wickremasinghe 2016 (18) | Systematic review of processes and tools for local decision-making in LMIC using information and evidence from health systems data | Search dates NR; published 2016; 14 databases searched | n=10 studies describing the approach of tools for decision-making; includes case studies and 1 realist evaluation. | Not explicitly summarized. Frameworks are reported to include prioritization, and estimates of budget and impact from local data. | This study focuses on decision-making in a specific context, using local data. Includes rather narrowly focused decision criteria |

### 2. EtD frameworks used by key organizations

Fourteen organizations that use EtD frameworks for recommendation formulation or decision-making were examined in detail (Table 2). One organization, the GRADE Working Group, is an informal network of individuals from a broad range of organizations, including academic institutions, national and international guideline development agencies, and healthcare provider organizations, among others.^20^ The GRADE Working Group does not, itself, publish guidelines, but rather develops processes and methods for use by other organizations which develop guidelines. The other thirteen organizations develop guidelines for specific audiences and with a clearly defined scope or set of topics: five are agencies of national governments,^21–25^ two are related to the World Health Organization (WHO),^26, 27^ one is a U.S. State agency (California Environmental Protection Agency (CalEPA)^28^) and the remainder are non-governmental organizations or academic groups.^2, 19, 29–31^

**Table 2.** Characteristics of key organizations. Evidence-to-decision frameworks: Characteristics of the organizations that developed the frameworks Abbreviations: EtD, evidence-to-decision; GRADE, Grading of Recommendations, Development and Evaluation; MCDA, multi-criteria decision analysis; NR, not reported; WHO, World Health Organization Footnotes: (^a^) Organization that developed the framework (^b^) Funder(s) for the development of the framework (^c^) Declaration of interests of the developers of the framework

| Table 2. Evidence-to-decision frameworks: Characteristics of the organizations that developed the frameworks |  |  |  |  |  |  |
| --- | --- | --- | --- | --- | --- | --- |
| Organization* | Type of organization<sup>a</sup> | Target audience;<br>Goal | Year established;<br>current version | Methods for development of the EtD<br>framework | Funder<sup>b</sup><br>Declarations of interest<sup>c</sup> | Use and sources of<br>evidence to inform EtD<br>criteria |
| 1 **Advisory Committee on Immunization Practices (ACIP) (US Centers for Disease Control and Prevention) (36)** | "US Federal advisory committee that provides expert advice to the Director of CDC and the Secretary of the US Department of Health and Human Services in the form of recommendations on the use of vaccines and related agents for control of vaccine-preventable diseases" | Public health programs in the US; health care providers and persons in the US civilian population;<br>To develop recommendations on how to use vaccines to control disease in the US | Established 1964; EtD framework updated June 2018 | Result of expert meeting Feb 2018; adapted from GRADE framework | NR<br>Authors report no conflicts of interest | Systematic reviews of the evidence on benefits and harms |
| 2 **Breast Cancer Prevention Partners (BCPP) (29)** | Non-governmental organization in the US | State policy-makers, health systems and healthcare providers;<br>Prevention of breast cancer; focus on the intersection of breast cancer and environmental health | Established 1992;<br>Methods published September 2020 | Expert committee, input from community representatives and other stakeholders | California Breast Cancer Research Program<br>NR | Systematic reviews of the evidence on interventions |
| 3 **California Environmental Protection Agency (CalEPA) (28)** | US state governmental agency | Policy-makers and regulators in the State of California, USA;<br>To restore, protect and enhance the environment, to ensure public health, environmental quality and economic vitality | Agency established 1991<br>A Guide to Pesticide Regulation, updated in 2017 | NR | NR<br>NR | Systematic reviews for risk assessment |

| Table 2. Evidence- |  |  |  |  |
| --- | --- | --- | --- | --- |
| Organization* | Assessment of the quality/certainty of the body of evidence | Names for recommendation or evaluation | No recommendation | Research or knowledge gaps |
| **Advisory Committee on Immunization Practices (ACIP) (US Centers for Disease Control and Prevention) (36)** | GRADE system | "Recommendations will be communicated in the framework in one of three categories: 1) ACIP recommends vaccination for all persons in an age group or a group at increased risk for vaccine-preventable disease; 2) ACIP does not recommend the use of a vaccine; or 3) the ACIP recommendation relies upon guidance of the clinician in the context of individual clinician-patient interactions to determine whether or not vaccination is appropriate for a specific patient." | "In some instances (e.g., when additional information is needed), ACIP might not make a recommendation, and this option is also reflected in the EtR framework separately." | "ACIP workgroups should identify research needs and, if appropriate, prioritize them. In formulating research needs, workgroups should be as specific as possible about what is needed and why. One format is EPICOT (ref: Brown P et al. BMJ 2006;333:804-6)." |
| **Breast Cancer Prevention Partners (BCPP) (29)** | NR | NR | NR | Research gaps highlighted for each risk factor |
| **California Environmental Protection Agency (CalEPA) (28)** | NR | NR | NR | Discusses authorization of agents in the context of research |

| Organization* | Type of organization<sup>a</sup> | Target audience;<br>Goal | Year established;<br>current version | Methods for development of the EtD<br>framework | Funder<sup>b</sup><br>Declarations of interest<sup>c</sup> | Use and sources of<br>evidence to inform EtD<br>criteria |
| --- | --- | --- | --- | --- | --- | --- |
| California Environmental Protection Agency (CalEPA) (37) | US state governmental agency | Alternative Analysis analysts, preparers, practitioners, and responsible entities;<br>To provide tools, information sources, and best practice approaches to help conduct Alternatives Analysis; challenges " responsible entities to reduce or eliminate toxic chemicals in the products consumers buy and use.... To identify Priority Products containing Chemicals of Concern, and for responsible entities to identify, evaluate, and adopt better alternatives." | Agency established 1991<br>Alternative Analysis Guide, version 1.1, July 2020 | NR | NR<br>NR | "Relevant factors" for alternatives analysis "can be quantified by available information or based on qualitative information" |
| 4 Evidence and Values Impact on Decision Making (EVIDEM) (30) | Developed by a group of authors at private and academic institutions | Variable depending on decision maker;<br>To provide a practical framework to facilitate decision making in a variety of contexts and to enhance the communication of decisions | Established 2006; first published 2008;<br>Current framework: 2018 | Review of the literature and or decision-making processes in use; identification of the steps and components of decision-making processes; framework developed by the study authors with input from thought leaders and stakeholders | "The publication costs for this article were funded by Mark O'Freil, the Brinson Foundation, and the Payne Family Foundation"; "No sources of funding were used to conduct this study and internal sources of support for the study were provided by the WSB and BioMedCom Consultants."<br>Authors declare no completing interests | Relevant evidence is collected and assessed |
| 5 Grading of Recommendations, Assessment, Development and Evaluation (GRADE) (clinical - individual or population perspective) (11, 16) | Consortium of academics and other guideline stakeholders | Varies with the organization/entity using the GRADE system;<br>To standardize assessment of the certainty (quality) of a body of evidence and the formulation of recommendations | Established 2000;<br>Methods continuously updated; EtD framework published 2016 | Started with GRADE Working Group approach (Guyatt 2008); iterative process; included brainstorming, feedback from stakeholders, application to recommendations and decisions, user testing (Alonso-Coello BMJ 2016.Introduction) | European Commission<br>Authors report no conflicts of interest | Systematic reviews, primary research, expert opinion; systematic review preferred depending on the criteria |

| Organization* | Assessment of the quality/certainty of the body of evidence | Names for recommendation or evaluation | No recommendation | Research or knowledge gaps |
| --- | --- | --- | --- | --- |
| California Environmental Protection Agency (CalEPA) (37) | Uncertainty analysis performed for individual factors assessed (sensitivity analysis or scenario analysis); no recommendation for quality of the body of evidence | NR | NR | NR |
| Evidence and Values Impact on DEcision Making (EVIDEM) (30) | "Quality of evidence" is assessed using bespoke tools based on existing tools, tailored to each type of evidence; sub-criteria include relevance, validity, completeness of reporting, type of evidence, and consistency. | Varies across end-users of this framework | NR | NR |
| Grading of Recommendations, Assessment, Development and Evaluation (GRADE) (clinical - individual or population perspective) (11, 16) | GRADE system | Strong, weak/conditional/discretionary, for or against the intervention | Possible when "pros and cons of the intervention or option and the comparison are so closely balanced that the panel is not prepared to make a weak recommendation in one direction or the other." and when "there is so much uncertainty that the panel concludes... that a recommendation would be speculative." | Rcommended when: i) there is insufficient evidence supporting an intervention for a guideline panel to recommend the intervention's use; ii) further research has a large potential for reducing uncertainty about the effects of the intervention; and iii) further research is deemed good value for the anticipated costs. |

| Organization* | Type of organization<sup>a</sup> | Target audience;<br>Goal | Year established;<br>current version | Methods for development of the EtD<br>framework | Funder<sup>b</sup><br>Declarations of interest<sup>c</sup> | Use and sources of<br>evidence to inform EtD<br>criteria |
| --- | --- | --- | --- | --- | --- | --- |
| **GRADE (coverage decisions)** (9) | Consortium of academics and other guideline stakeholders | Third-party payers (public or private) for the purpose of deciding whether and how much to pay for drugs, tests, devices or services and under what conditions;<br>To standardize assessment of the certainty (quality) of a body of evidence and the formulation of recommendations | Established 2000;<br>Methods continuously updated; EtD framework published 2017 | Iterative process: brainstorming workshops, consultation with advisory group, user testing, feedback, application to different types of coverage decisions | European Commission<br>One author reports having received funding from the pharmaceutical industry; all other authors report no conflicts of interest | Systematic reviews, primary research, expert opinion; systematic review preferred depending on the criteria |
| **GRADE (health system and public health decisions)** (6) | Consortium of academics and other guideline stakeholders | Population or health system; specific population perspective depends on the nature of the decision; e.g., could be societal or governmental;<br>To standardize assessment of the certainty (quality) of a body of evidence and the formulation of recommendations | Established 2000;<br>Methods continuously updated; EtD framework published 2018 | Iterative process based on the GRADE clinical EtD: brainstorming workshops, consultation with stakeholders, survey of policy-makers, experience with policy briefs, applied the framework to examples, conducted workshops, observed guideline panels using the framework, conducted user testing | European Commission<br>Authors report no conflicts of interest | Research evidence ("information derived from studies that used systematic and explicit methods"); "additional considerations include other evidence such as routinely collected data, and assumptions and logic" |
| **6 Guide Community Preventive Services (US Centers for Disease Control and Prevention)** (24, 34) | Independent body of experts, funded by the US government and supported by the US Centers for Disease Control and Prevention | Policy-makers at the state or community level, or in community or healthcare organizations, businesses, or schools;<br>To improve health or prevent disease | Established 1998; Methods updated 2017 (unpublished) | Review of the US Preventive Services Task Force methods, input from experts in systematic reviews, the Task Force, and other external advisors | NR<br>NR | Systematic reviews of benefits and harms |
| **7 Institute for Clinical and Economic Review (ICER)** (31) | Independent, non-profit research organization based in the US | Health system managers, policy makers, payers<br>"evaluates medical evidence and convenes public deliberative bodies to help stakeholders interpret and apply evidence to improve patient outcomes and control costs." | Established 2006;<br>Methods updated Oct. 2020 | The initial framework was developed with input from a multi-stakeholder workgroup; followed by national public comment, review and feedback by a broad range of stakeholders. | ICER (which receives its funding from government grants and non-profits foundations); a separate policy program is funded in part by health insurers and other industries<br>NR | Systematic reviews of comparative effectiveness |

| Organization* | Assessment of the quality/certainty of the body of evidence | Names for recommendation or evaluation | No recommendation | Research or knowledge gaps |
| --- | --- | --- | --- | --- |
| **GRADE (coverage decisions) (9)** | GRADE system | Not covering, coverage only in the context of research, covering with price negotiation, restricted coverage, and full coverage | GRADE clinical EtD guidance likely applies | GRADE clinical EtD guidance likely applies |
| **GRADE (health system and public health decisions) (6)** | GRADE | Strong, weak/conditional/discretionary, for or against the intervention | GRADE clinical EtD guidance likely applies | GRADE clinical EtD guidance likely applies |
| **Guide Community Preventive Services (US Centers for Disease Control and Prevention) (24, 34)** | Strong, sufficient, insufficient strength of evidence based on quality of execution (study limitations), suitability of study design, number of studies, consistency, meaningfulness of effect size | Recommend, recommend against, insufficient evidence | Yes, "insufficient evidence", i.e. unable to determine effectiveness | Each chapter includes research and knowledge gaps focusing on effectiveness, applicability in other populations, economic consequences, implementation barriers, and opportunities to improve technical efficiency |
| **Institute for Clinical and Economic Review (ICER) (31)** | Systematic reviews of comparative effectiveness include an assessment of certainty of the body of evidence. | Provide assessment as to an intervention's "value for money". | NR | Consider future research needs |

|  | Organization* | Type of organization<sup>a</sup> | Target audience;<br>Goal | Year established;<br>current version | Methods for development of the EtD<br>framework | Funder<sup>b</sup><br>Declarations of interest<sup>c</sup> | Use and sources of<br>evidence to inform EtD<br>criteria |
| --- | --- | --- | --- | --- | --- | --- | --- |
| 8 | **International Society for Pharmacoeconomics and Outcomes Research (ISPOR)**<br>(19, 40, 42) | Non-profit, multidisciplinary, multistakeholder professional organization in pharmacoeconomics and outcomes research | Variable depending on who is making decisions<br>To assess the value of new technologies (value is defined from an economic perspective and includes "gross value" (what individuals or others acting on their behalf would be willing to pay to acquire more health care or other goods or services), and "opportunity cost" (what benefits or other resources they are willing to forgo to obtain them)) | Established 1995;<br>2018 | Task force appointed by ISPOR, with input from advisory board and stakeholder panel; reviewed existing examples of value assessment frameworks; through discussion and with feedback and review, arrived at final framework . | NR<br>NR | Data are used to measure the performance of alternatives; sources include systematic reviews, modelling, expert opinion, and other approaches as appropriate. |
| 9 | **National Institute for Health and Care Excellence (NICE); focus on guidelines**<br>(25, 35) | UK government organization; a non-departmental public body that provides national guidance and advice to improve health and social care in England. | Individual healthcare providers; local authorities, commissioners and managers; other providers of health and social services;<br>To produce evidence-based recommendations on a range of topics, including prevention and management of specific conditions, improving health, managing medicines, providing social care and support, and planning services for communities | Established 1999<br>Methods guidance published 2014, updated October 2018 | "The processes and methods described in this manual are based on internationally recognized standards, and the experience and expertise of the teams at NICE, the contractors..., NICE committee members and stakeholders. They are based on internationally accepted criteria of quality...and primary methodological research and evaluation undertaken by the NICE teams. They draw on the Guideline Implementability Appraisal tool to ensure that recommendations are clear and unambiguous, making them easier to implement." | NR<br>NR | Systematic reviews of the evidence; "colloquial evidence" can be included (e.g. expert testimony); for economic analyses: systematic review of existing models; may perform de novo model |

| Organization* | Assessment of the quality/certainty of the body of evidence | Names for recommendation or evaluation | No recommendation | Research or knowledge gaps |
| --- | --- | --- | --- | --- |
| **International Society for Pharmacoeconomics and Outcomes Research (ISPOR)**<br>(19, 40, 42) | MDCA includes an "uncertainty analysis to understand the level of robustness of the MCDA results" | MDCA results in an assessment of the "total value" of the alternatives under consideration. | NR | NR |
| **National Institute for Health and Care Excellence (NICE); focus on guidelines**<br>(25, 35) | Individual study quality assessed according to study design; the certainty or confidence in the findings should be presented at outcome level using GRADE or GRADE-CERQual; body of evidence for each outcome is high, moderate, low, very low "certainty or confidence of evidence"; it integrates a review of the quality of cost-effectiveness studies... it does not use 'overall summary' labels for the quality of the evidence across all outcomes: "strength of evidence (reflecting the appropriateness of the study design to answer the question and the quality, quantity and consistency of evidence)": classified as no, weak, moderate, strong or inconsistent evidence | NICE uses the wording of recommendations to reflect the strength of the evidence (e.g. "offer, advise, refer versus consider") | "If evidence of efficacy or effectiveness for an intervention is either lacking or too low quality for firm conclusions to be reached, the committee.... may: make a 'consider' recommendation based on the limited evidence... decide not to make a recommendation and make a recommendation for research... recommend that the intervention is used only in the context of research... recommend not to offer the intervention." | Include recommendations for research; "The committee should select up to 5 key recommendations for research that are likely to inform future decision-making (based on a systematic assessment of gaps in the current evidence base)." |

|  | Organization* | Type of organization<sup>a</sup> | Target audience;<br>Goal | Year established;<br>current version | Methods for development of the EtD<br>framework | Funder<sup>b</sup><br>Declarations of interest<sup>c</sup> | Use and sources of<br>evidence to inform EtD<br>criteria |
| --- | --- | --- | --- | --- | --- | --- | --- |
| 10 | **Navigation Guide** (2, 15) | Non-profit collaboration between governmental and non-governmental (including academic) organizations in the US and Europe | Clinicians, policy-makers, professional societies, health care organizations, government agencies making prevention-oriented guidelines<br>To provide a methodology for evaluating the evidence and to support evidence-based decision-making in environmental health | Published 2011<br>Current version 2014 | Collaborative process among clinicians, systematic review and guidelines experts, statistics, epidemiology, and environmental health scientists; based on GRADE | For 2009–2013, support for the development and dissemination of the Navigation Guide methodology was provided by the Clarence Heller Foundation, the Passport Foundation, the Forsythia Foundation, the Johnson Family Foundation, the Heinz Endowments, the Fred Gellert Foundation, the Rose Foundation, Kaiser Permanente, the New York Community Trust, the Philip R. Lee Institute for Health Policy Studies, the Planned Parenthood Federation of America, the National Institute of Environmental Health Sciences ... and U.S. EPA STAR grants.<br>Authors report no conflicts of interest. | Systematic reviews of the evidence on the risks to human health of exposure to chemicals, and the effects of prevention and mitigating interventions |
| 11 | **Scottish Intercollegiate Guideline Network (SIGN)** (21, 32) | Supported by the Scottish government, but with editorial independence | Health and social care professionals, patients;<br>To understand and use medical evidence to make decisions about healthcare, reduce unwarranted variations in practice, make sure patients get the best care available, improve healthcare across Scotland | Established 1993; Handbook first published 2008; Current version: November 2019 | Based on 2013 GRADE/DECIDE work | Core funding for SIGN activities comes from Healthcare Improvement Scotland<br>NR | Systematic reviews of the evidence |

| Organization* | Assessment of the quality/certainty of the body of evidence | Names for recommendation or evaluation | No recommendation | Research or knowledge gaps |
| --- | --- | --- | --- | --- |
| Navigation Guide (2, 15) | The quality of individual studies and the overall body of evidence is rated, including for human and animal data | NR<br>(Statements about the health risks of substances include: known to be toxic, probably toxic, possibly toxic, not classifiable, or probably not toxic.) | NR | NR |
| Scottish Intercollegiate Guideline Network (SIGN) (21, 32) | GRADE system | Strong recommendation against; conditional recommendation against; recommendation for research and possibility conditional recommendation for use restricted to trials; conditional recommendation for; strong recommendation for | NR | Include recommendations for research |

| Organization* | Type of organization<sup>a</sup> | Target audience;<br>Goal | Year established;<br>current version | Methods for development of the EtD<br>framework | Funder<sup>b</sup><br>Declarations of interest<sup>c</sup> | Use and sources of<br>evidence to inform EtD<br>criteria |
| --- | --- | --- | --- | --- | --- | --- |
| 12 **US Preventive Services Task Force** (22, 33) | Independent body of experts, funded by the US government | Primary care clinicians, also policy-makers, payers, patients;<br>Tp provide recommendations for preventive care for general, primary care populations in the US who are asymptomatic with respect to the condition addressed by the intervention | Established 1984; Procedure manual Dec 2015 | Developed by the Methods Working Group of the USPSTF using an iterative process based on the methods literature, international standards and practices; approved by the Task Force | NR<br>NR | Systematic reviews of benefits and harms; sometimes on contextual questions also |
| 13 **World Health Organization (WHO) - guidelines** (26) | United Nations agency | National and local policy makers and program managers;<br>To prevent disease and promote health | Established 1948; Handbook for guideline development, 2nd edition: 2014 | Adopted directly from the then-current (2014) GRADE approach | The Bill & Melinda Gates Foundation<br>NR | Systematic reviews of benefits and harms and other considerations as indicated |
| 14 **WHO-INTEGRATE** (27) | Developed for WHO; applies to any entity making public health or health system guidelines | National and local policy makers and program managers;<br>To prevent disease and promote health | Published 2019 | i) an analysis of WHO's norms and values; ii) a systematic review of EtD criteria in clinical care and public health; iii) key informant interviews; iv) application to completed WHO guidelines; v) focus groups; vi) peer review; and vii) the development of guidance and prompts for completing the EtD. | Funding provided by the World Health Organization Department of Maternal, Newborn, Child and Adolescent Health through grants received from the United States Agency for International Development and the Norwegian Agency for Development Cooperation<br>One author is a WHO employee; two authors are members of the GRADE Working Group | Evidence gathered as needed to inform key EtD considerations; systematic reviews for key criteria |

| Organization* | Assessment of the quality/certainty of the body of evidence | Names for recommendation or evaluation | No recommendation | Research or knowledge gaps |
| --- | --- | --- | --- | --- |
| **US Preventive Services Task Force**<br>(22, 33) | Assessment of certainty across the analytic framework, where certainty is "the likelihood that the USPSTF assessment of the net benefit of a preventive service is correct"; "assessing the certainty of evidence requires a complex synthesis of all evidence across the entire analytic framework" in order to determine if "the results observed in the individual studies in the body of evidence would be expected when the intervention is delivered to asymptomatic persons by providers in US primary care settings". | Grades (or strength) of recommendations:<br>A. The USPSTF recommends the service. There is high certainty that the net benefit is substantial.<br>B The USPSTF recommends the service. There is high certainty that the net benefit is moderate, or there is moderate certainty that the net benefit is moderate to substantial.<br>C. The USPSTF recommends selectively offering or providing this service to individual patients based on professional judgment and patient preferences. There is at least moderate certainty that the net benefit is small.<br>D. The USPSTF recommends against the service. There is moderate or high certainty that the service has no net benefit or that the harms outweigh the benefits. | I Statement. The USPSTF concludes that the current evidence is insufficient to assess the balance of benefits and harms of the service.<br>Evidence is lacking, of poor quality, or conflicting, and the balance of benefits and harms cannot be determined. | Reports on evidence gaps for each clinical preventive service the Task Force reviews; and there is an annual report to congress that focuses on evidence gaps as well |
| **World Health Organization (WHO) - guidelines** (26) | GRADE system | Strong, conditional, for or against | No explicit guidance provided; not prohibited | Added as a requirement in 2019; specific methods and guidance under development |
| **WHO-INTEGRATE**<br>(27) | No specific guidance provided but an assessment is recommended | No specific guidance provided | NR | NR |

Two organizations focus primarily on clinical care: the Scottish Intercollegiate Guideline Network (SIGN)^32^ and the US Preventive Service Task Force (USPSTF).^33^ The Guide to Community Preventive Services (GCPS) focuses on interventions aimed at groups, communities or health systems^34^ and WHO and WHO-INTEGRATE primarily on public health interventions.^26, 27^ Three organizations are oriented to health technologies, with a prominent focus on economic evaluations and resource considerations.^19, 30, 31^ The UK National Institutes for Health and Care Excellence (NICE) examines a broad range of clinical, public health, and social interventions.^35^ The Advisory Committee on Immunization Practices (ACIP) focuses exclusively on vaccine recommendations for U.S. populations.^36^ Breast Cancer Prevention Partners (BCPP),^29^ CalEPA,^28, 37^ and the Navigation Guide^2, 15^ focus on the human health effects of hazardous substances in the environment.

The amount of information available on each of the 14 key organizations varied greatly. There are a large number of publications describing the GRADE system for guideline development, including the GRADE EtD framework and its use.^20^ On the other hand, there is only one document describing the approach of the BCPP.^29^

#### Development process and expertise

For the organizations that developed *de novo* EtD frameworks and described the process for developing them, all used an iterative approach based on an examination of other organizations, with input from experts in guideline methods and evidence synthesis (Table 2). The most comprehensive approach was taken by Rehfuess and colleagues^27^ in development of the WHO-INTEGRATE framework. Their approach included development of a theoretical framework, a review of WHO basic documents, a systematic review of EtD criteria, input from a range of stakeholders, and a thematic analysis to identify key domains.^38^

It was difficult to discern the expertise of contributors to framework development; most groups appeared to consist mainly of academic, generalist guideline methodologists with either clinical or public health experience. Social scientists led work on WHO-INTEGRATE^27^ and contributed to the GRADE public health framework.^6^ Eight of the 14 organizations reported who funded the development of the EtD framework and five reported the conflicts of interest of the framework developers; the remaining organizations did not provide this information.

#### Decision criteria

Eighteen frameworks were examined in detail: one from each of the 14 organizations, except for CalEPA and GRADE, where two^28,37^ and four ^6, 9, 11, 16^ unique frameworks were examined, respectively. There were significant commonalities across frameworks in the criteria for formulating recommendations (Table 3 and Figure 1). Unsurprisingly, all included consideration of benefits, except the CalEPA framework for pesticides^28^ which examines risks (of environmental exposures) and not benefits. All frameworks included an assessment of harms of the intervention under consideration. Fifteen of the 18 frameworks included some assessment of certainty or quality of the body of evidence across outcomes in the decision-making process (in contrast to an assessment of individual outcomes as part of an evidence review discussed below): BCPP, one of the CalEPA frameworks, and ISPOR (International Society for Pharmacoeconomics and Outcomes Research) did not.^19, 28, 29^ Some measure of costs, resource use or cost-effectiveness was found in all frameworks except BCPP, USPSTF and the GCPS.^29, 33, 34^ For the latter two frameworks, the U.S. government prohibits consideration of financial resources in the decision-making process. Other decision criteria were variably included: feasibility (13 frameworks) equity (12), values (11), and acceptability (11). Only two frameworks included human rights: WHO^26^ and WHO-INTEGRATE frameworks.^38^

**Figure 1.**
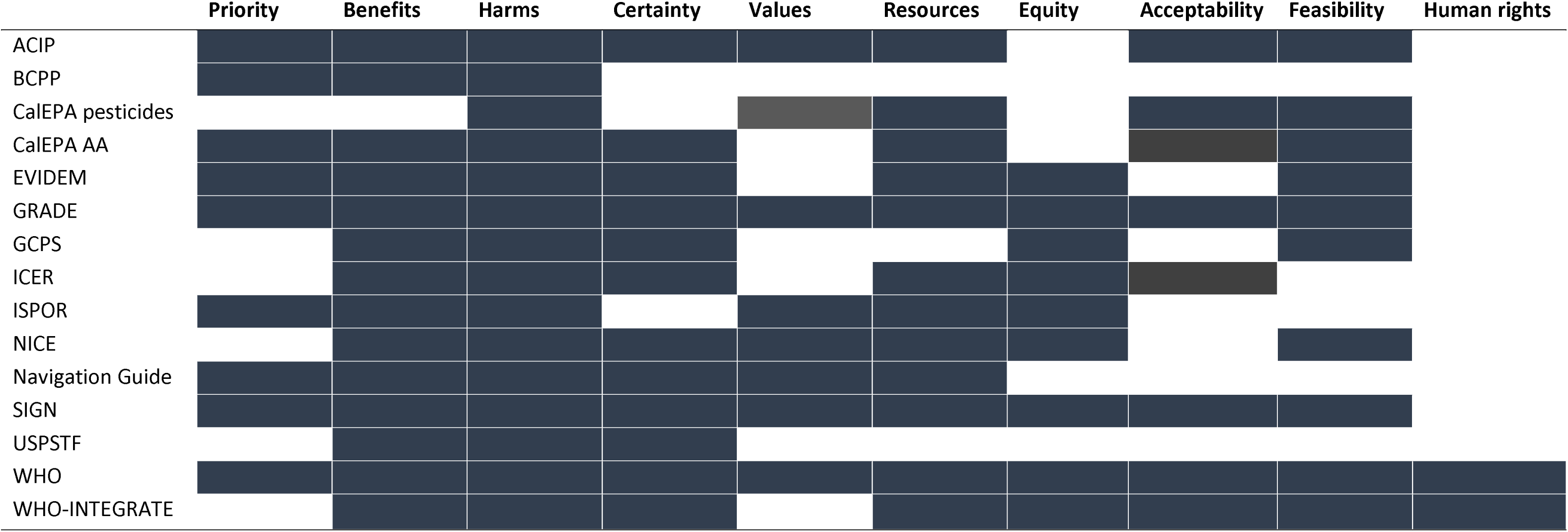
Evidence-to-decision criteria for each key organization. **Legend**. Across the top of the figure, broad categories of evidence-to-decision criteria are presented. For each listed organization, the cell is shaded if their evidence-to-decision framework encompasses one or more criteria within a category. Abbreviations: AA, Alternatives Analysis ACIP, Advisory Committee on Immunization Practices BCPP, Breast Cancer Prevention Partners CalEPA, California Environmental Protection Agency EVIDEM, Evidence and Values Impact on DEcision Making GRADE, Grading of Recommendations, Assessment, Development and Evaluation GCPS, Guide Community Preventive Services ICER, Institute for Clinical and Economic Review ISPOR, International Society for Pharmacoeconomics and Outcomes Research NICE, National Institutes for Health and Care Excellence WHO, World Health Organization

**Table 3.**
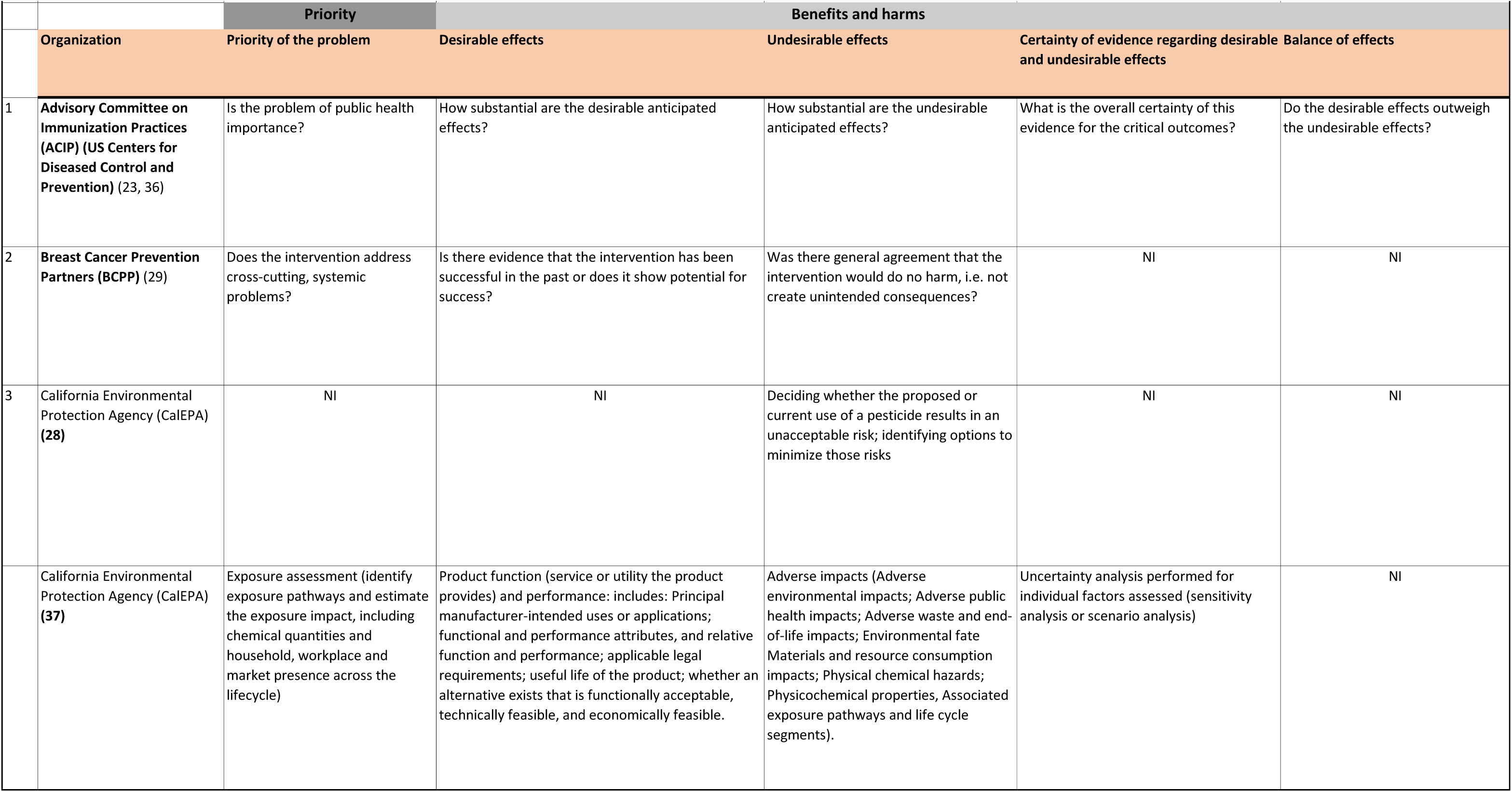
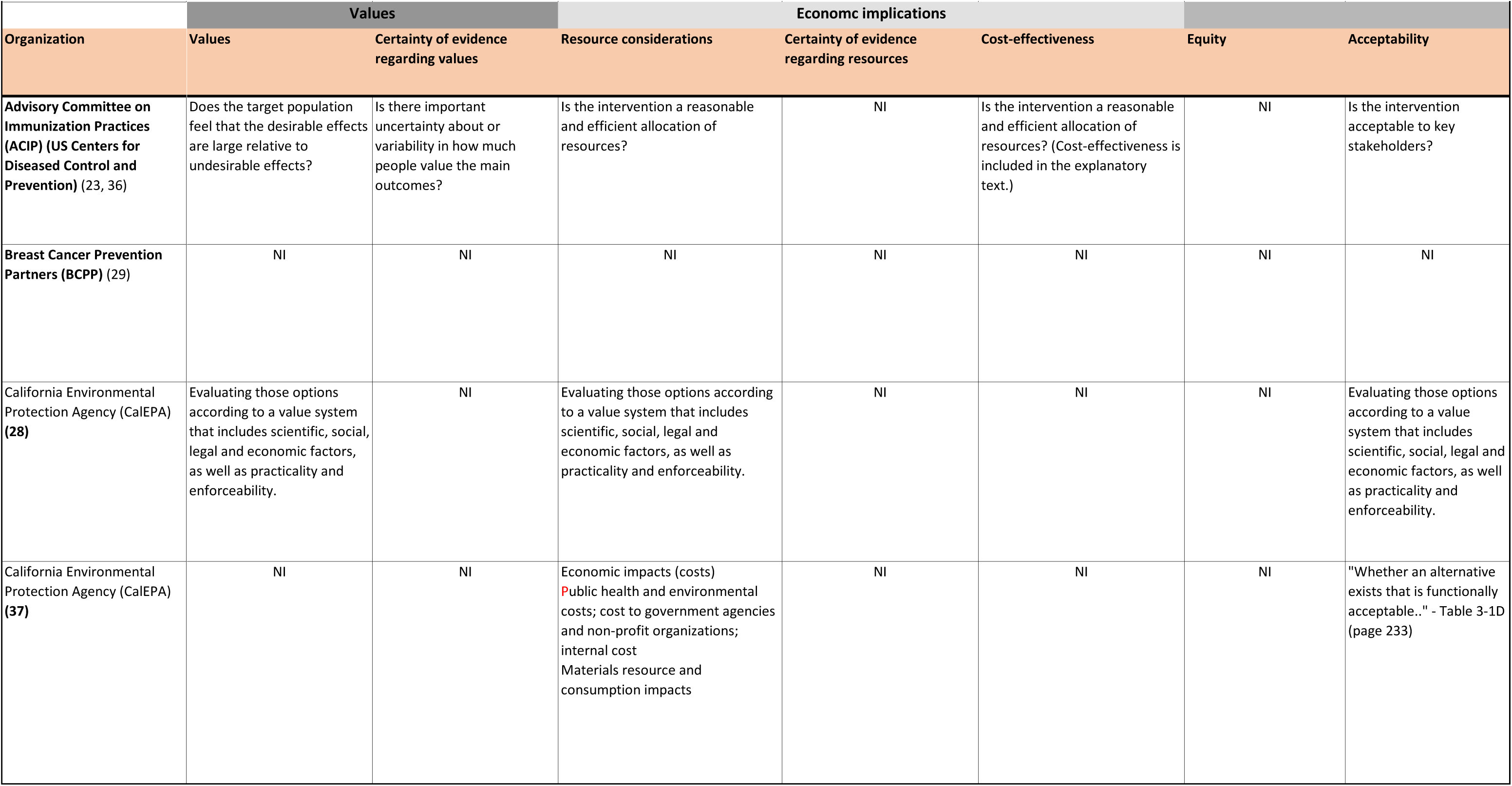
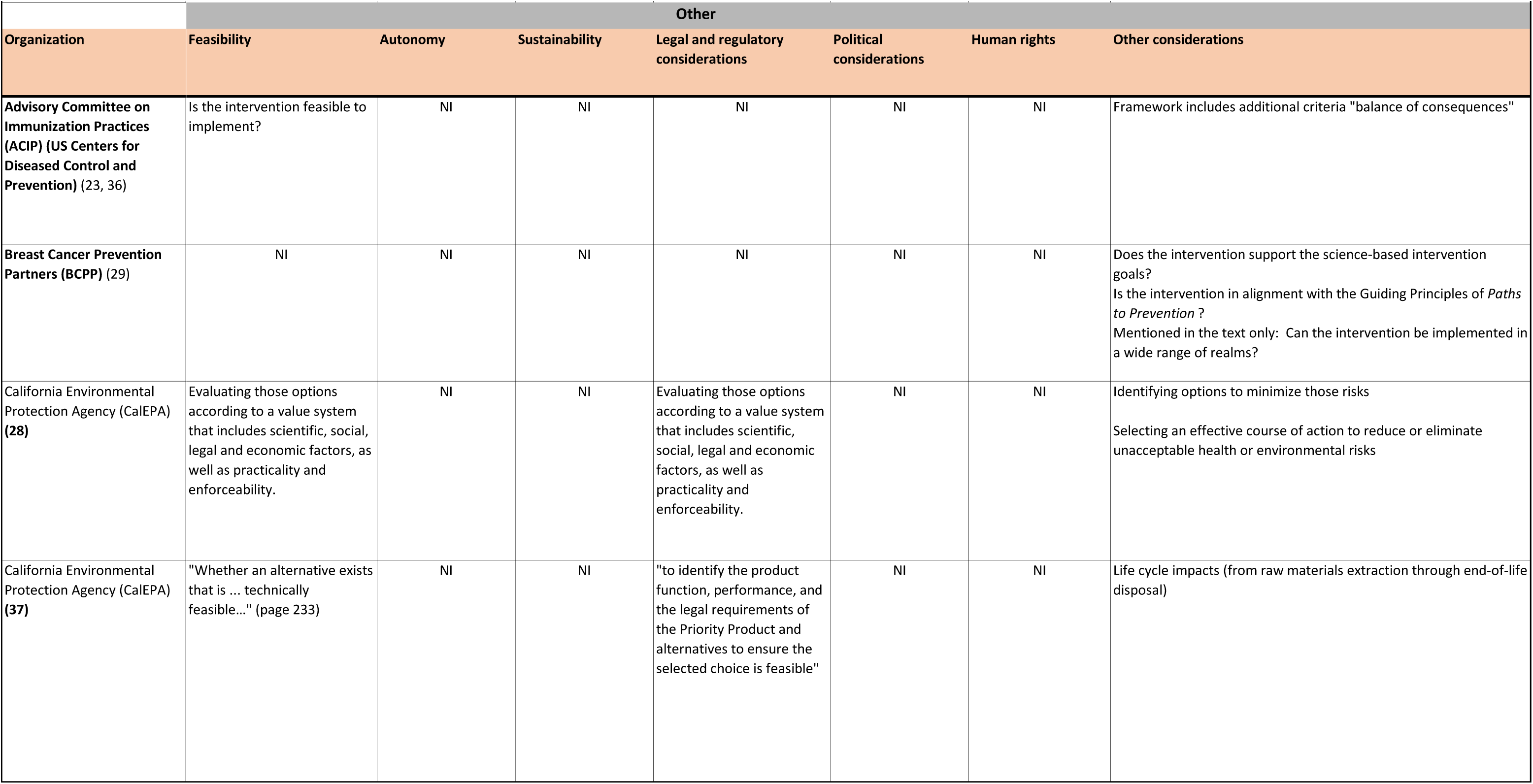
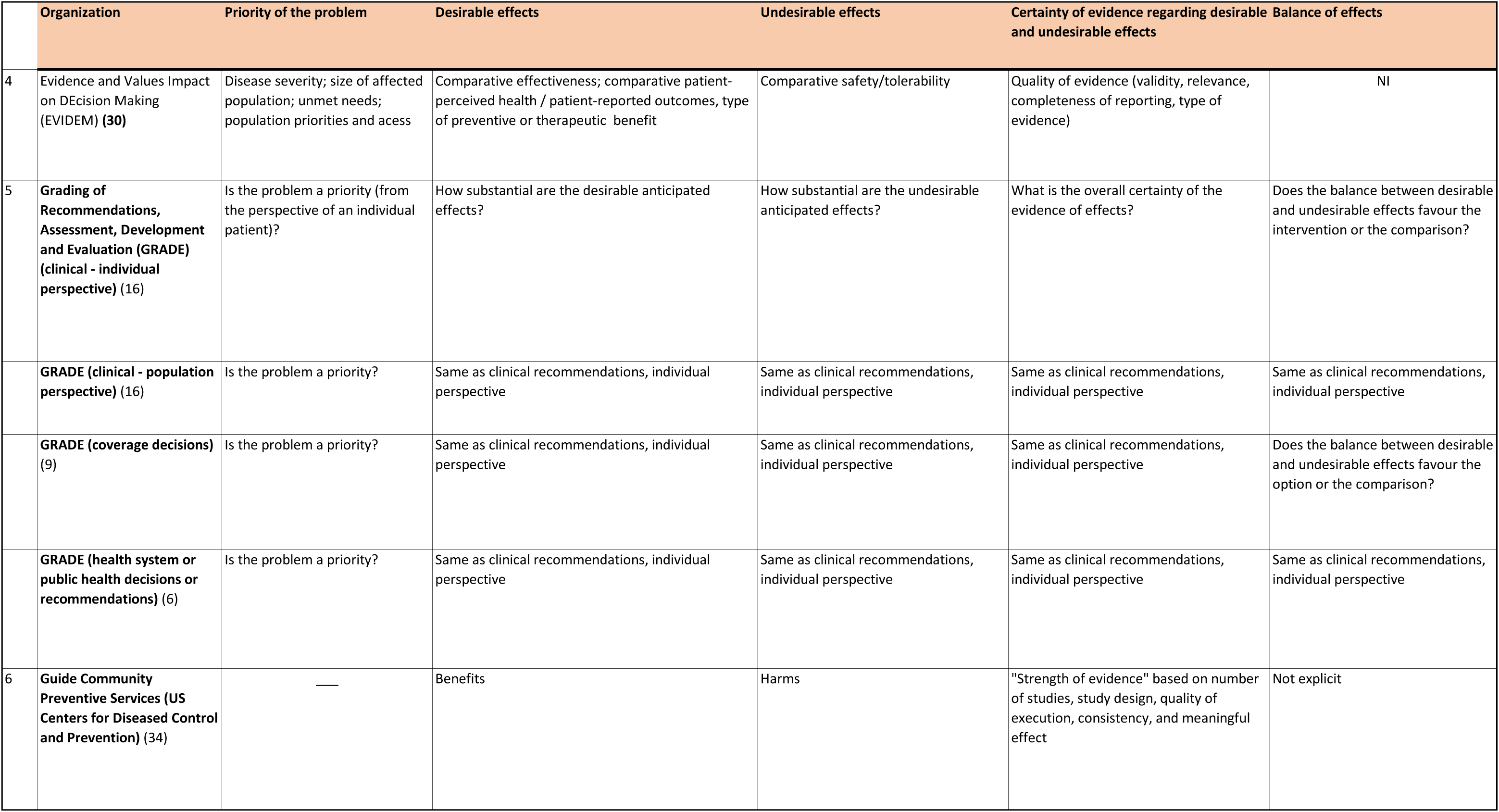
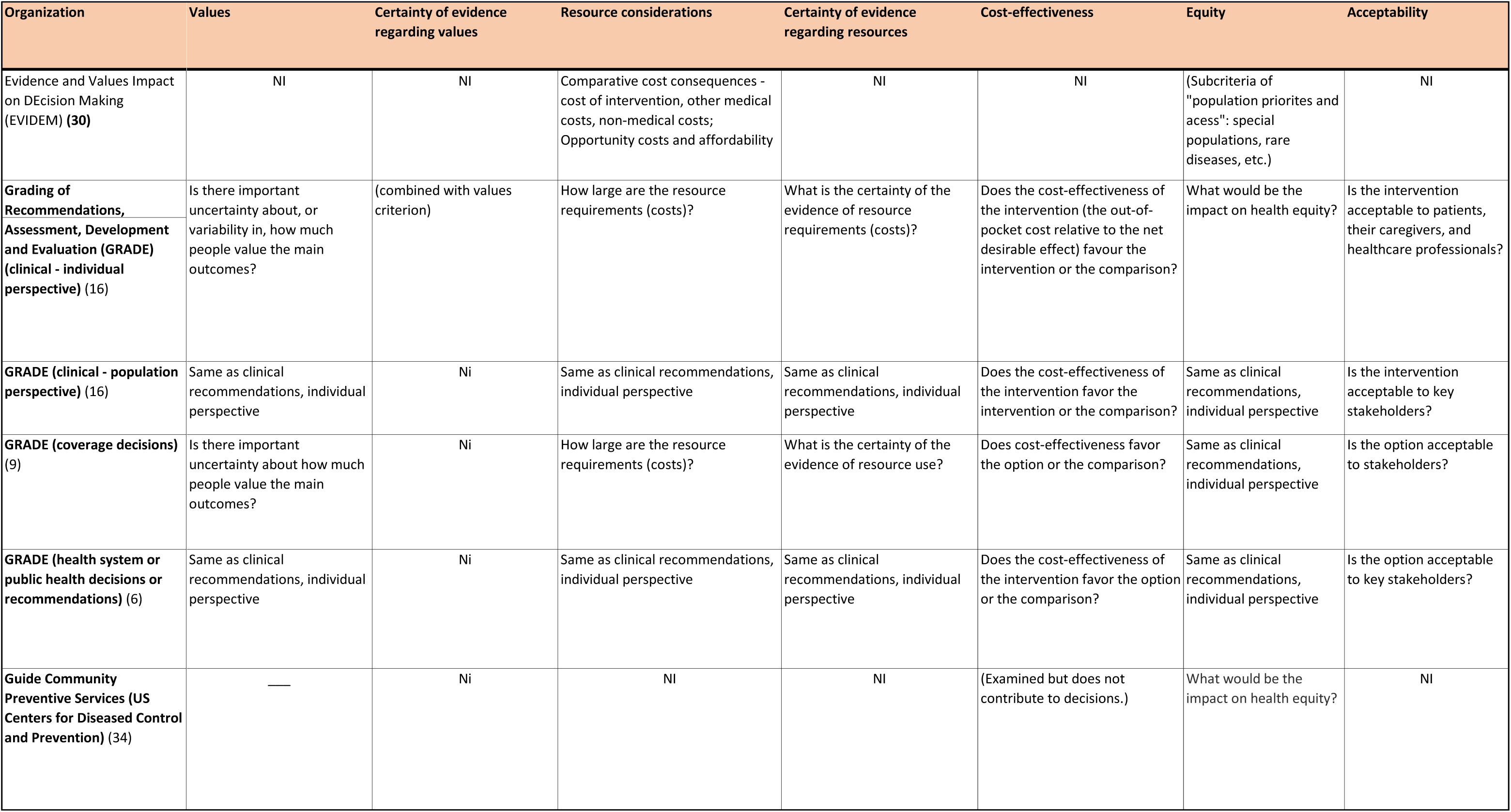
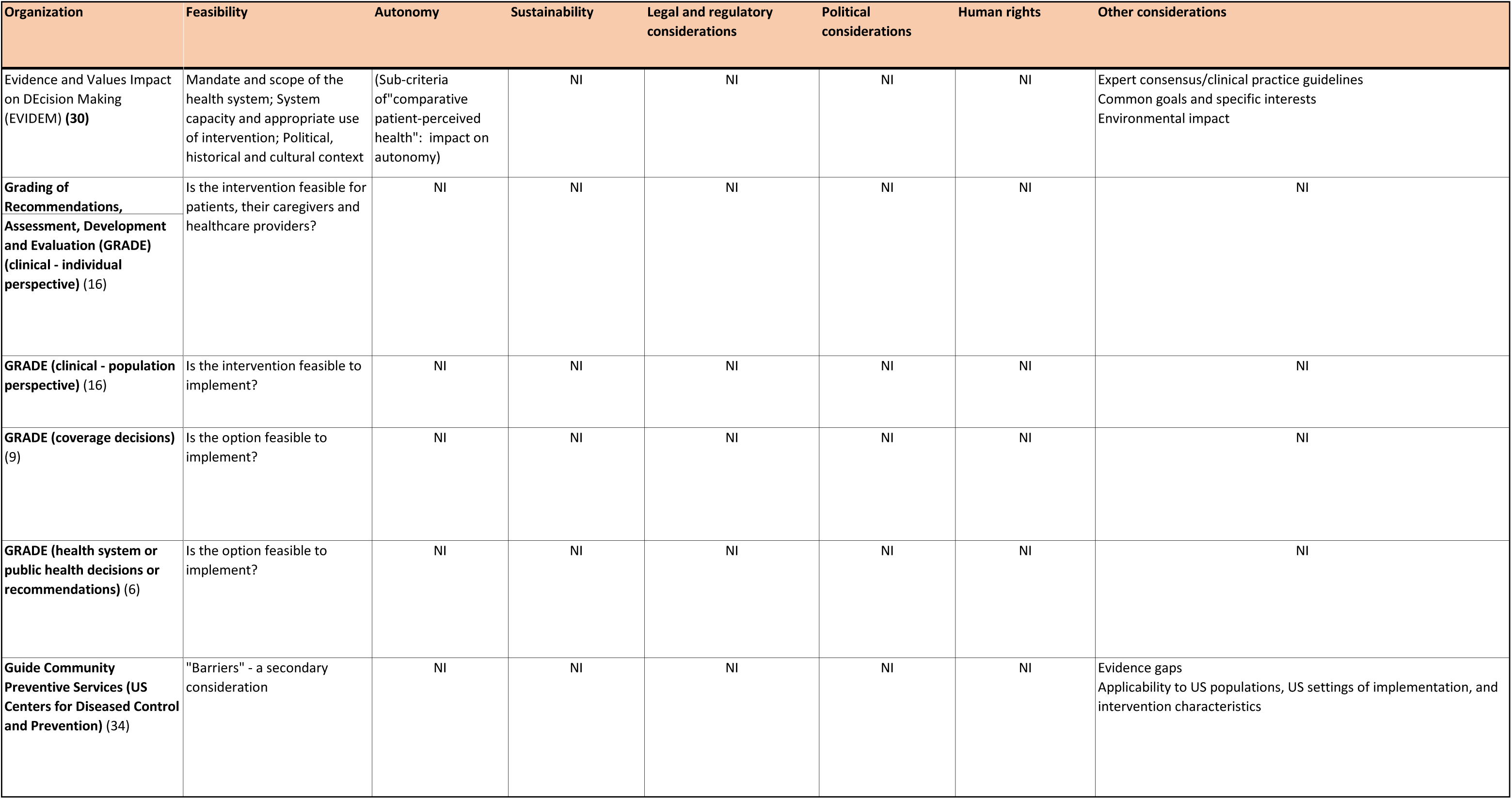
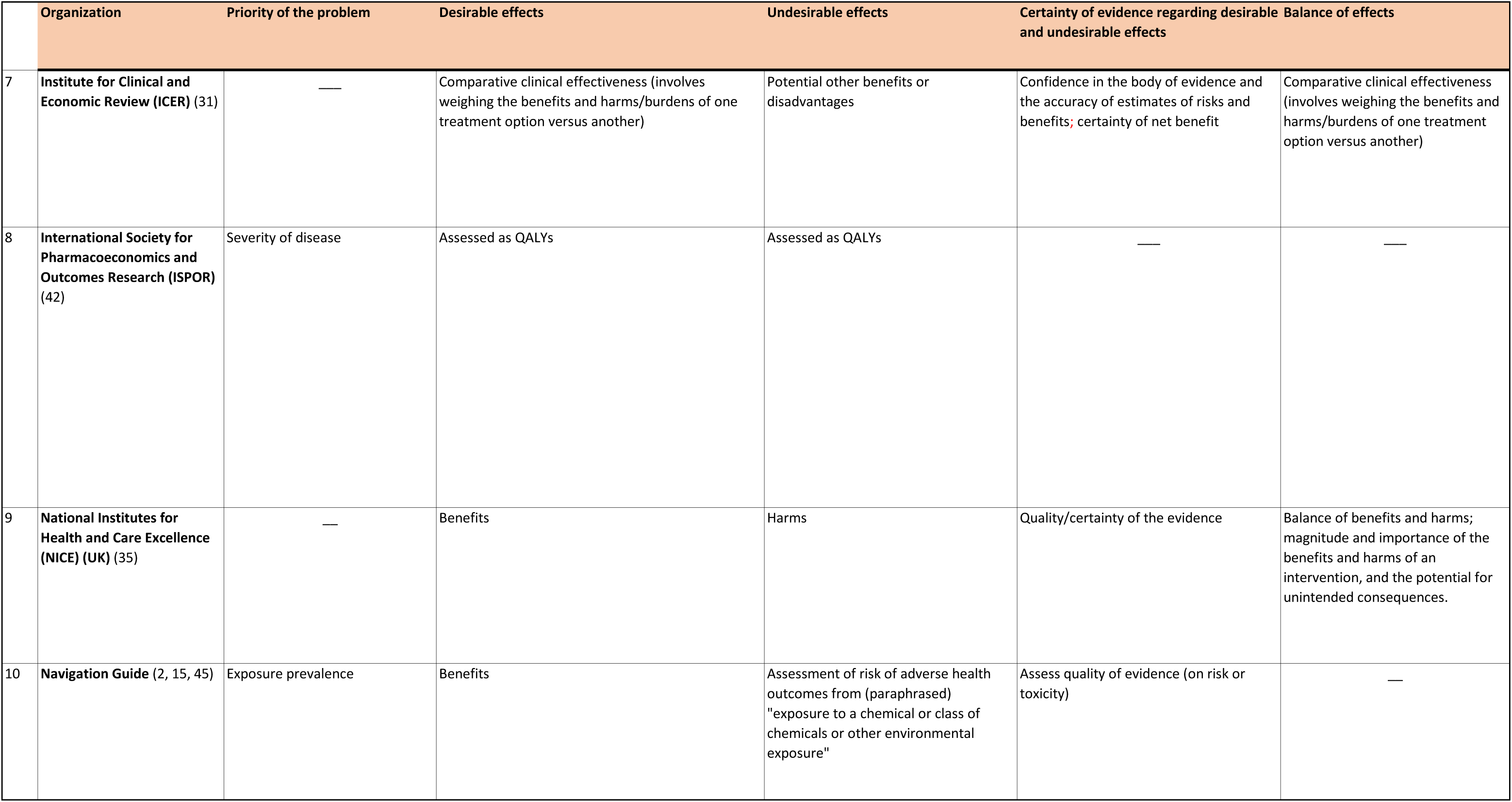
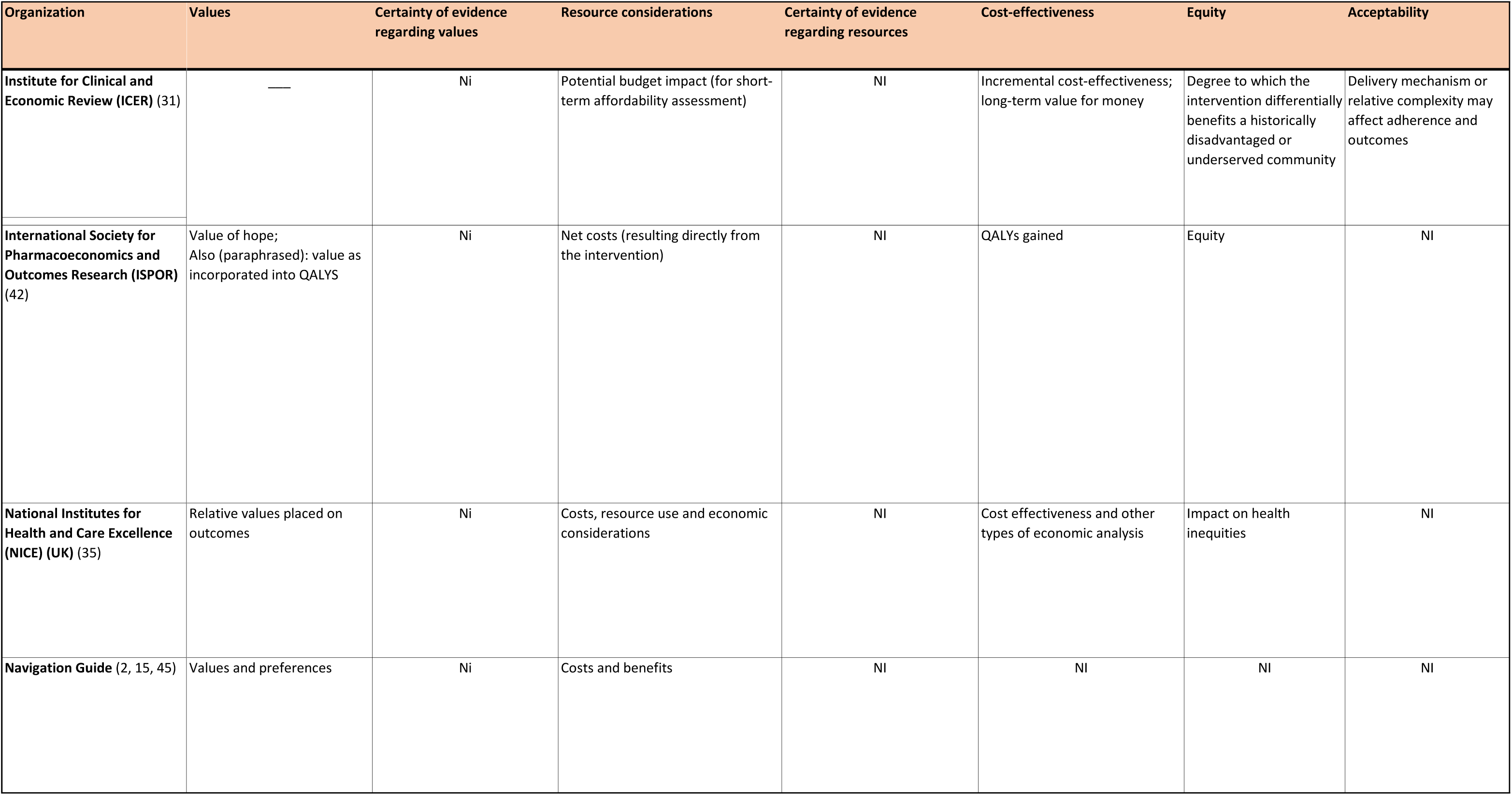
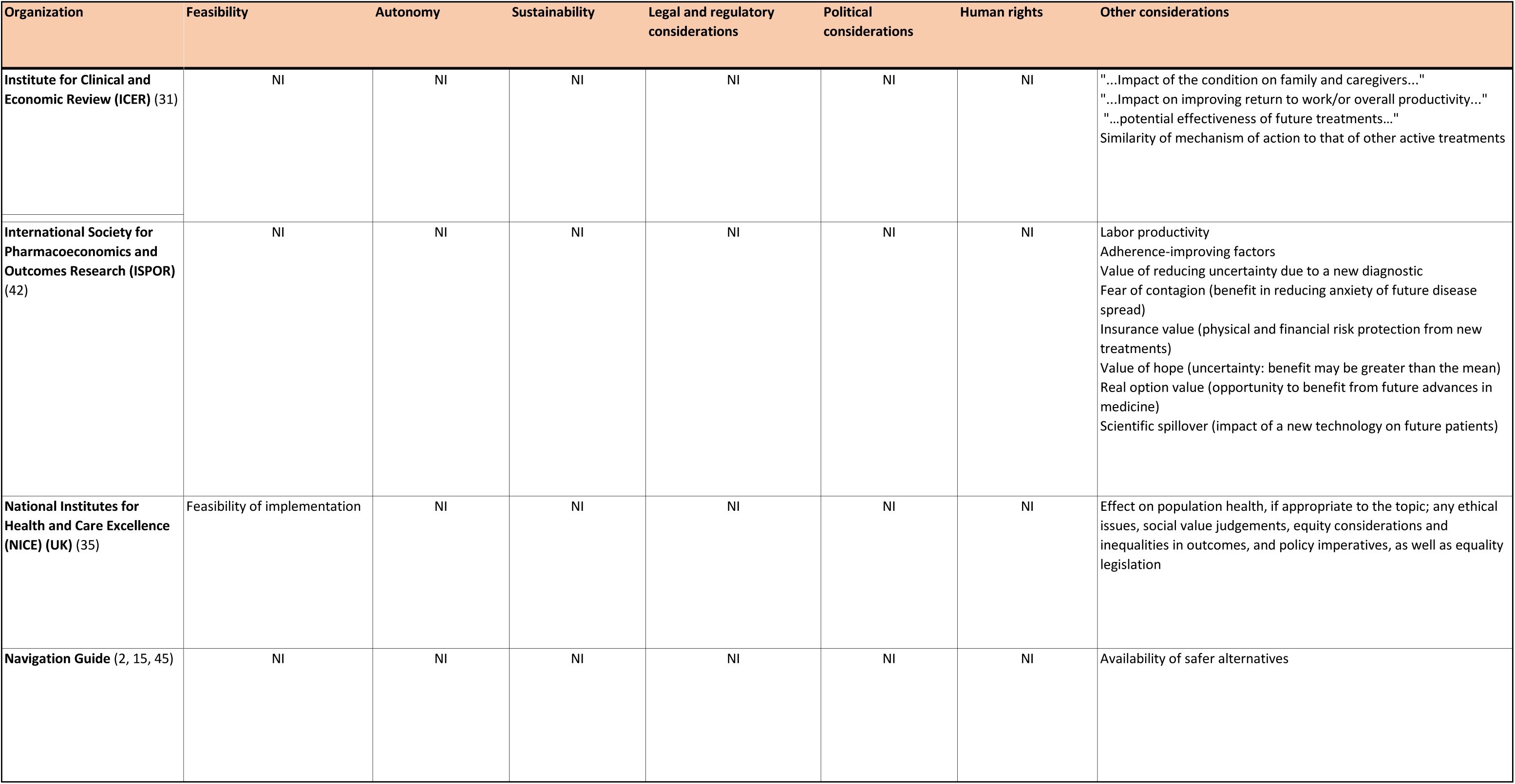
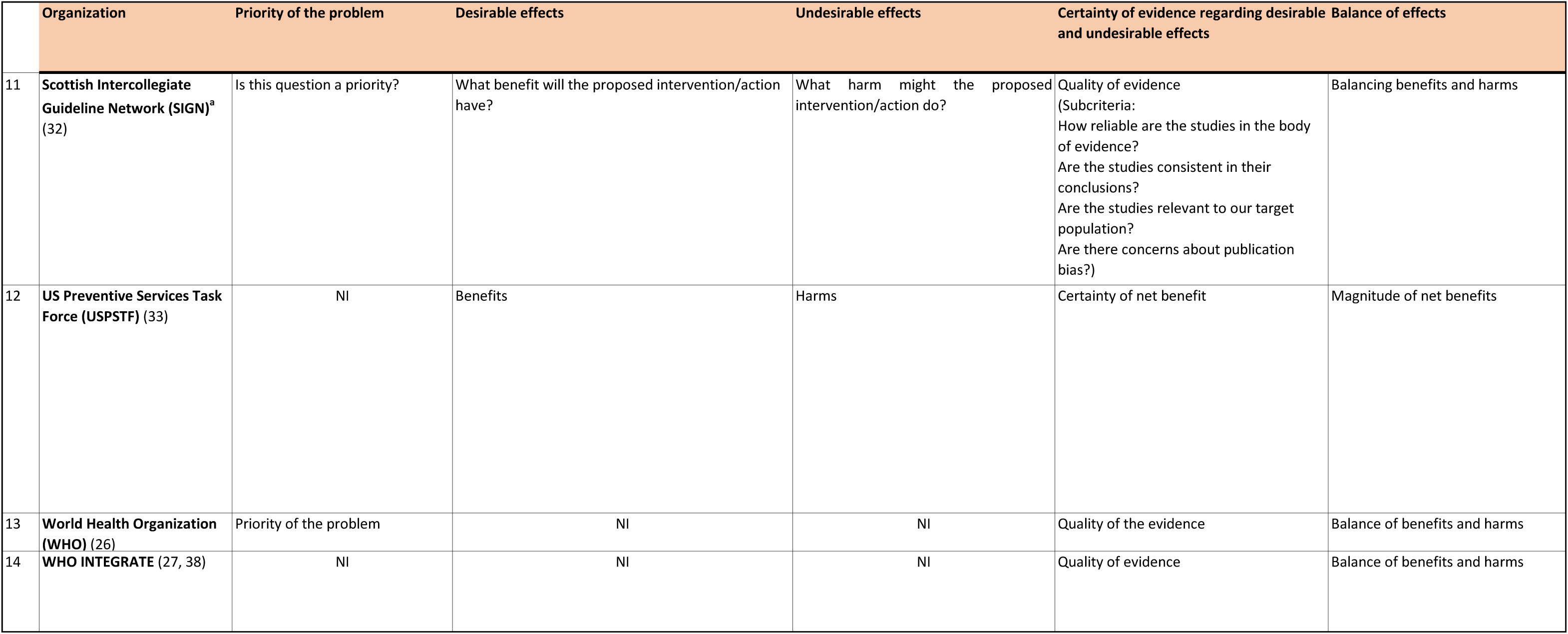
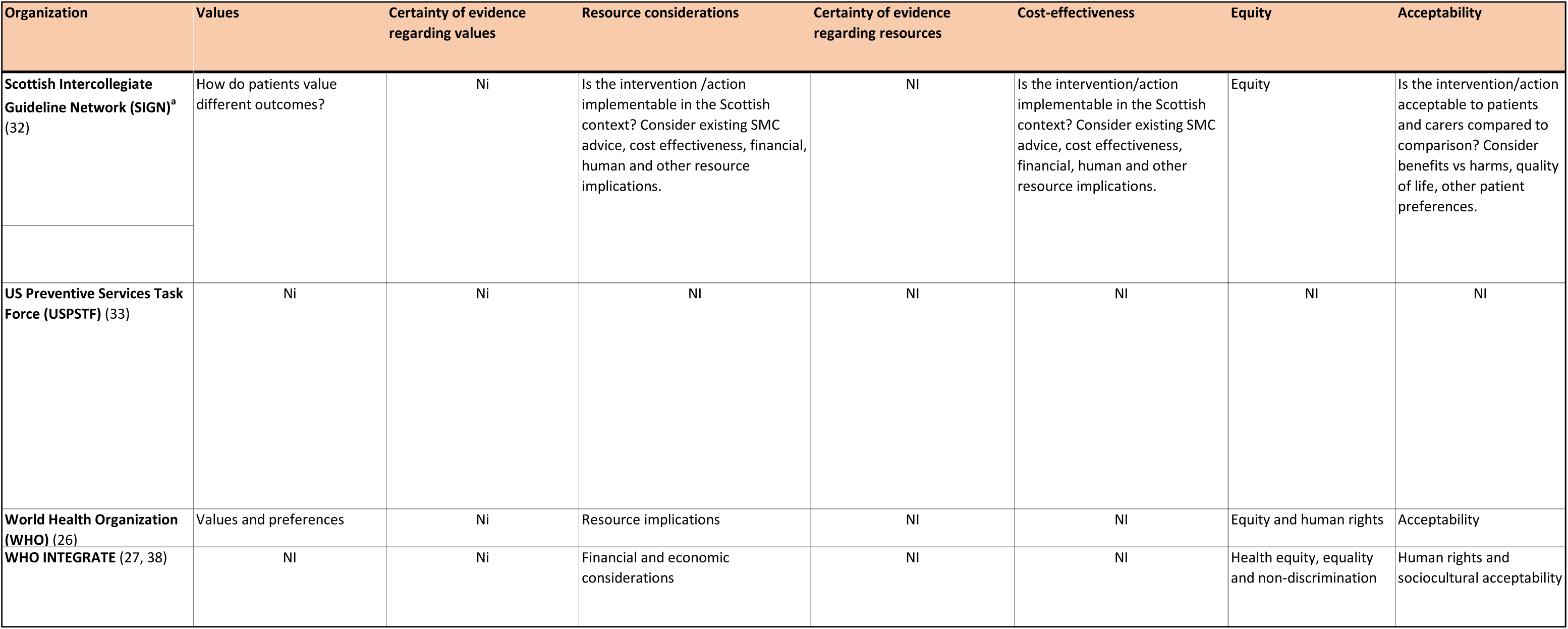
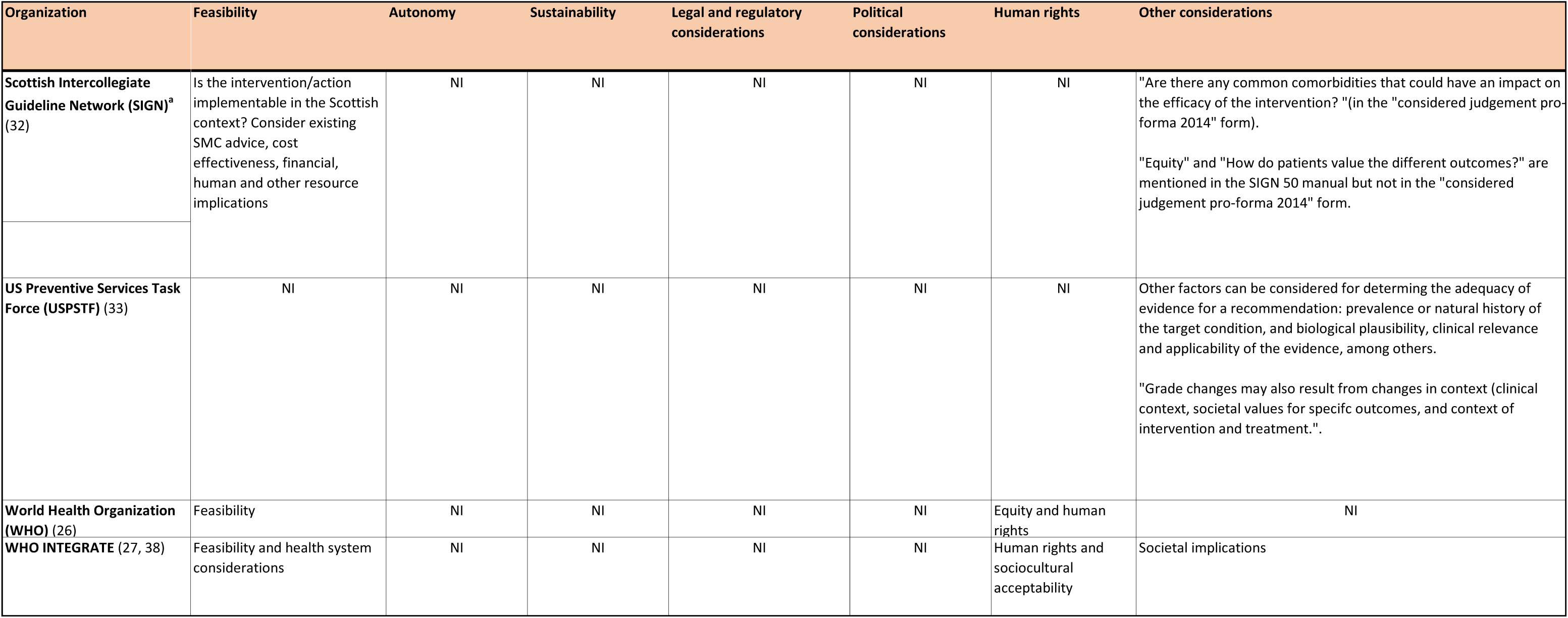
Evidence-to-decision criteria for the key organizations. Criteria included in evidence-to-decision frameworks Abbreviations: NI, not included Footnotes (^a^) The criteria listed here are from both the “Considered judgement pro-forma 2014” form and the headings in the SIGN 50 guideline handbook as these differ.

“Priority of the problem” was included in the frameworks based on GRADE,^26, 32, 36^ as well as the BCPP,^29^ EVIDEM (Evidence and Values Impact on DEcision Making),^30^ ISPOR^19^ and Navigation Guide frameworks.^15^ However, in the background materials for several other frameworks, the burden of the disease (number of affected persons, severity of the disease, or economic and social costs) was mentioned as an important consideration when prioritizing an intervention for development of a guideline or policy. This latter consideration, however, differs from use of this criterion when deciding on the strength and direction of a recommendation.

The meaning of the criterion “values and preferences” has varied over time and across publications. This phrase encompasses two rather different constructs: the relative value that persons affected by the recommendations place on the outcomes of the intervention (values), and the preferences such persons have regarding the intervention options. The GRADE Working Group initially combined these two constructs^5^ with a definition encompassing both.^39^ The 2016 update of the GRADE EtD framework distinguished these constructs, with the inclusion of two separate criteria: variability of the value affected persons place on the main outcomes, and acceptability of the intervention^11^ to various stakeholders. The WHO EtD framework, published in 2014,^26^ preceded the GRADE modifications of 2016,^11^ includes “values and preferences” as well as “acceptability” and “feasibility”. The text explains that “values and preferences” “*pertain to the relative importance people assign to the outcomes associated with the intervention or exposure; they have nothing to do with what people think about the intervention itself*.”^26^ Other key organizations include a decision criterion related to values using verbiage closely aligned with GRADE^15^ and focused exclusively on the relative value of outcomes,^32, 35^ or they include the concept but with a somewhat different meaning.^19, 28, 36^

#### Evidence used to inform each criterion when organizations make decisions

All of the frameworks report that the EtD criteria should be informed by evidence obtained from a variety of sources, with an emphasis on systematic reviews of research evidence (Table 2). Twelve of the organizations recommend an assessment of the quality (validity or certainty) of the body of evidence for important outcomes as part of the evidence review, and most recommend the GRADE system with or without modifications. Only BCPP^29^ and one of the CalEPA guidelines,^28^ do not recommend some assessment of quality or certainty of the evidence as part of the systematic review process. The approaches used by the GCPS ^34^ and USPSTF^33^ differ somewhat from GRADE in their assessment of the “strength of evidence” and “certainty of net benefit”, respectively. The EtD frameworks with an economic focus describe varied approaches to assessing quality of the body of evidence.^19, 30, 31^

#### Nomenclature for recommendations

Nine of the included organizations provide specific guidance on the nomenclature for the types of recommendations formulated based on the EtD criteria (Table 2). The most common categorization of recommendations among those nine organizations was two levels of strength both for and against a recommendation, i.e., four categories.^11, 26, 32^ GRADE has extensive documents on this issue and uses the terms “strong, weak or conditional“.^5^ The USPSTF has five categories of recommendations,^33^ while ACIP^36^ and the GCPS^34^ have three. The economic-focused frameworks refer to “value” in various ways.^30, 31, 40^ NICE uses the wording of recommendations to reflect the strength of the evidence (e.g., offer, advise, consider) rather than standardized terms to represent the strength of the recommendation.^35^

Five organizations provide explicit guidance on the situations where recommendations cannot be formulated due to insufficient evidence or a close balance between benefits and harms (Table 2).^11, 33–36^ The USPSTF provides the most detailed guidance on this situation.^41^ Ten of the organizations suggest including knowledge or research gaps with the recommendations, with particular emphasis in ACIP, NICE, GCPS and the USPSTF.

#### Specific evidence-to-decision frameworks

##### i GRADE evidence-to-decision frameworks

GRADE includes EtD frameworks for four different purposes: i) clinical recommendations, individual perspective; ii) clinical recommendations, population perspective; iii) coverage decisions; and iv) health system and public health recommendations/decisions. (There is a fifth GRADE EtD framework for diagnostic, screening and other tests but this was not included in this review of interventions considered most relevant for application to environmental health.^11^) These frameworks are all very similar, all with 12 criteria covering the same concepts, with some variation in verbiage, tailored to the different audiences (Table 3). There is more emphasis on resource considerations, equity, acceptability and feasibility for health systems and public health decisions than for individual patient clinical recommendations.^6, 9^

##### ii Evidence-to-decision frameworks based directly on GRADE

ACIP’s EtD framework is derived directly from GRADE,^36^ while NICE^25^ and SIGN’s^32^ EtD criteria closely resemble those of GRADE. WHO uses the GRADE EtD framework which was current when the *WHO Handbook for Guideline Development*, 2^nd^ edition was published in 2014.^26^ Guideline development groups supported by WHO can modify this EtD framework to meet the needs of individual guidelines, and are encouraged to use the current GRADE EtD framework published in 2016.^11, 16^

##### iii Other evidence-to-decision frameworks

The WHO-INTEGRATE EtD framework version 1.0,^27, 38^ first published in 2019 (Table 2), was developed in response to a perceived need to take a complexity perspective into account and to incorporate public health and WHO-specific values when developing WHO guidelines. The developers also sought to address several weaknesses that they noted in the GRADE EtD:^27^ the absence of a theoretical framework; failure to adequately consider the role of social and economic determinants of health; inadequate consideration of complex interventions, system changes, and the environment in which interventions are implemented; and an undue focus on benefits and harms and not on other considerations critical for decision-making in public health.

WHO-INTEGRATE includes six broad criteria (Table 3): balance of benefits and harms; human rights and sociocultural acceptability; health equity, equality and non-discrimination; societal implications; financial and economic considerations; and feasibility and health system considerations.^27^ A seventh criterion, quality of evidence, is a meta-criterion that applies to each of the six other criteria. WHO-INTEGRATE also includes a number of sub-criteria. For example, the main criterion “Health equity, equality and non-discrimination”^27^ includes several sub-criteria related to the intervention: impact on health equality and/or health equity, distribution of benefits and harms, affordability, and accessibility.

One of the key concepts of the WHO-INTEGRATE approach is the careful consideration of the relative importance of the various EtD criteria at the planning stage of guideline development. Logic models or conceptual frameworks are strongly recommended in order to understand the interrelationships between the intervention and the context in which it is delivered, with prioritization of EtD considerations for the specific decision or recommendation. Relevant evidence is then sought on this subset of criteria.

The USPSTF^22^ develops an analytic framework depicting the causal pathway by which the intervention may achieve its effects (benefits and harms) and the various questions which must be researched to address the overarching question of net benefit of the intervention. The USPSTF’s recommendations are based primarily on net benefits and the certainty thereof, although other factors may be considered (Table 3).

The GCPS modeled their original processes and methods on those of the USPSTF^34^ with some later modifications (personal communication Dr David Hopkins, 25 January 2021). Based on systematic reviews focused mainly on benefits and harms of the intervention, the beneficial effects of the intervention are assessed as strong, sufficient or insufficient strength of evidence. This assessment is then “upgraded” or “downgraded” based on factors including harms, equity considerations, magnitude of effect and applicability of the evidence to US populations.

Three of the frameworks from key organizations reflect an economic perspective for decision-making in health systems or in decisions on coverage at a system or national level: EVIDEM,^30^ ISPOR,^42^and ICER (Institute for Clinical and Economic Review).^31^ These frameworks all included consideration of comparative effectiveness. ISPOR presents data as quality adjusted life years (QALYs) and net costs, and includes other considerations such as “value of hope” and “insurance value” (Table 3).^42^ ICER^31^ focuses on comparative clinical effectiveness and incremental cost-effectiveness along with “contextual considerations”, such as “improving return to work and/or overall productivity”. EVIDEM^30^ also focuses on comparative effectiveness, along with cost consequences and other considerations (e.g., “environmental impact”).

##### iv Evidence-to-decision frameworks in environmental health

The three frameworks which focus on interventions in environmental health were generally less well developed and lacked specificity compared with the clinical and public health frameworks.^2, 28, 29^ BCPP developed a comprehensive breast cancer primary prevention plan for the US State of California, focusing on primary prevention and systemic interventions to reduce the incidence of breast cancer, based on the principles of social justice and equity.^29^ This organization prioritized a large number of potential interventions using criteria focused on alignment with the organization’s goals and guiding principles, whether the intervention addresses cross-cutting or systemic problems, past success of the intervention, and potential harms.

CalEPA’s Office of Environmental Health Hazard Assessment (OEHHA) leads California’s assessment of health risks posed by environmental contaminants^43^ and works with the State Department of Pesticide Regulation (DRP) to develop regulations related to pesticide worker health and safety. In response to a review by the US National Academy of Sciences (NAS),^44^ CalEPA published an update of their methods for risk assessment and risk management of pesticides^28^ which focuses on an assessment of a range of options and their risks, and examination of scientific, social, legal, and economic factors, as well as practicality and enforceability.

CalEPA’s Department of Toxic Substances Control Safer Products and Workplaces Program has published a formal approach to alternative analysis of potentially hazardous components of consumer products.^37^ This approach guides decisions on product removal, redesign or other change to reduce the potential impact of the chemical of concern. The analysis compares alternative products across factors including potential chemical exposures, public and environmental health impact, product function and requirements, and economic implications. This analysis considers all relevant life cycle consequences of the chemical or product of concern, from inputs into manufacturing, through end-of-life disposal. While not referenced in the summary “List of Factors for Consideration in Alternatives Analysis” (Appendix 3-1),^37^ other considerations are mentioned in the document text. For example, the entity performing the analysis determines which impacts are most significant based on its values. Likewise, the variable impacts across sensitive subpopulations are to be considered with respect to exposure to or use of the product. Feasibility is also discussed in the text, with respect to the assessment of product function, performance, and the legal requirements.^37^ Finally, acceptability is considered in regards to the function of alternative compounds.

The Program on Reproductive Health and the Environment at the University of California at San Francisco has developed a process and methods for integrating scientific findings from human and nonhuman studies to determine the overall strength of the evidence on the effects of environmental exposures on outcomes in humans.^45^ This approach, published as *The Navigation Guide*^2, 15, 45^ has adapted the GRADE approach to integrate multiple streams of evidence and a range of study designs, to draw conclusions on the quality and strength of the body of evidence of a substance’s toxicity. This assessment of toxicity is then combined with information on exposure (level, duration and timing) to provide an assessment of risk of adverse health outcomes. This risk assessment could then be combined with consideration of: 1) whether a less toxic agent is available as an alternative; and 2) “values and preferences”, to arrive at a recommendation.^2, 15^

## Discussion

A number of EtD frameworks have been developed in a variety of clinical, and public and environmental health disciplines, and there is significant overlap in the criteria used to inform recommendations or decisions across these frameworks. Benefits and harms are almost universally included, and an assessment of certainty or quality of the body of evidence and some measure of resource use were included in most frameworks examined. Other decision criteria such as values, equity, feasibility, acceptability were variably included, while only two frameworks encompassed human rights. There was variation across frameworks in terminology, definitions and presentation of EtD criteria. The five systematic reviews of EtD frameworks describe decision criteria that were similar to those presented in the frameworks from key organizations.

The 18 frameworks identified for 14 key organizations focused mainly on clinical medicine or public health interventions. Of these, only one was based on a conceptual framework, and rarely was a systematic review of potential criteria performed during the framework’s development. The GRADE framework is the most well developed, with extensive information and guidance on its methods. A number of the key organizations examined have adopted GRADE, often with minor modifications.^15, 26, 32, 35, 36^ Three of the four frameworks related to environmental health lacked detail on the specific criteria for decision-making.^19, 30, 31^ Frameworks originating in the HTA realm unsurprisingly focus on economic considerations, while also including criteria similar to those encompassed by other frameworks.

### General aspects of evidence-to-decision frameworks

The process and methods for developing EtD frameworks were often superficial and poorly reported. Only WHO-INTEGRATE^38^ and the GRADE coverage^8^ frameworks were developed following a systematic review of existing frameworks or potential decision criteria. Only WHO-INTEGRATE describes a conceptual framework^27^ underpinning the EtD criteria. The lack of a conceptual framework represents a significant weakness in most EtD frameworks: key considerations may be missed and long-standing criteria may be perpetuated without adequate scrutiny.

GRADE has dominated guideline methods in health care and public health over the last 15 years. This has led to productive collaborations, standardization of processes and methods, and the development of packages of detailed methods and guidance by diverse contributors from across the globe. On the other hand, there are instances where intellectual dominance by a single group – i.e., monopolies of knowledge – can suppress innovation and slow down development processes.^46^

EtD frameworks largely focus on, and are optimal for, single-component interventions with simple linear pathways from intervention to outcomes. In addition, most EtD frameworks do not consider the context in which the intervention is delivered. With the exception of WHO-INTEGRATE, EtD frameworks do not explicitly or even implicitly incorporate a complexity perspective including the inter-relationship between the intervention and the context or environment in which it is delivered. Only the GCPS, the USPSTF, and WHO-INTEGRATE recommend the use of visual depictions of the relationships among the intervention components, the causal pathway, and the context for the intervention (i.e., analytic frameworks, logic models, or similar depictions). Such approaches facilitate examination of multiple key questions linked across the causal pathway for intervention effects, as well as the consideration of contextual factors and externalities which may be important for decision-making.

Guidance on if and when to identify, synthesize and integrate evidence on criteria other than benefits and harms of the intervention into the decision-making process was provided only for WHO-INTEGRATE.^27^ Most guideline development processes focus almost exclusively on benefits and harms in the evidence review and in discussions, without explicit efforts to prioritize other criteria or seek evidence to inform them. Qualitative evidence, particularly on considerations such as acceptability or feasibility, is rarely gathered. Meetings at which recommendations are formulated often include only brief, *ad hoc* discussions of these other criteria: this is a major limitation of most current approaches to recommendation formulation.

### Evidence-to-decision criteria across frameworks

It was difficult to compare specific EtD criteria across frameworks due to significant variation in approaches to lumping and splitting criteria across frameworks, as well as in the terminology and definitions used. While most frameworks presented broad categories of criteria (e.g., “equity”), only WHO-INTEGRATE^27^ provides detailed sub-criteria to facilitate understanding and decision-making. Some main criteria included several constructs: aggregating diverse elements within a single criterion can be difficult to implement in the decision-making context and when reporting the rationale for the recommendation or decision. For example, WHO-INTEGRATE’s “Human rights and sociocultural acceptability” includes both human rights and what other frameworks refer to as “preferences” or “acceptability”. CalEPA^28^ articulates in one question a group of criteria: “Evaluating those options according to a value system that includes scientific, social, legal and economic factors, as well as practicality and enforceability”, without providing additional guidance or sub-criteria for decision-makers.

There is also variability in how criteria are defined and whether operational definitions or guidance are provided. The criterion “values and preferences” is particularly problematic. Unfortunately, guideline developers continue to use the phrase “values and preferences”, often with unclear and variable meaning. The WHO EtD framework, published in 2014,^26^ includes an outdated definition of “values and preferences” which differs both from the 2008 GRADE guidance^5^ and from the 2016 modifications:^11, 16^ it needs to be updated.

The GRADE criterion of “priority of the problem” is also problematic. It was initially included in the GRADE framework to facilitate prioritization across interventions, such as at the national or sub-national levels. However, in most guideline development scenarios, the problem on which interventions and comparators are focused has already been determined to be of high priority. Thus, examination of the burden of disease and other priority considerations is irrelevant at the stage of recommendation formulation.

The criteria related to resource use vary considerably, and may include cost, affordability, infrastructure needs, personnel training, and measures of economic efficiency (e.g., cost-benefit and cost-effectiveness). This variability is due in part to the different perspectives of the sponsors of the EtD framework or of the target audiences for the organization’s products. For example, the USPSTF is prohibited by national legislation from considering cost or cost-effectiveness in their recommendations. On the other hand, for the UK’s NICE, cost-effectiveness is a critical part of their decisions. There is also variability in the extent and specificity of the guidance on how to incorporate resource use into decision-making. While well-developed for NICE,^35^ and reasonably so for GRADE,^47^ guidance is almost completely lacking for WHO^26^ and WHO-INTEGRATE.^27^

The criterion of “equity” is poorly described and little operational guidance is provided across the frameworks except for WHO-INTEGRATE which provides several sub-criteria for consideration under the main criterion of “health equity, equality and non-discrimination”.^27^

### Group decision making

Group decision making when formulating recommendations in guidelines is rarely a simple, linear process. While EtD frameworks support the normative aspect of decision making in terms of how expert panels ‘‘should’’ or ‘‘ought to’’ develop recommendations by providing a structure for decision making,^48–50^ many factors affect how groups make decisions.^48^ In addition to the features or characteristics of the decision itself (e.g., criteria in the EtD framework such as resource use and equity), other factors may play a role: i) situational or contextual factors (e.g., time pressure, social context, gender bias, political pressures); ii) individual characteristics of the decision maker (e.g., role on the expert panels, cultural and professional background, race/ethnicity, methodological expertise); and iii) individual panel member’s emotions and experiences (e.g., personal experiences with a disease). How these various factors contribute to decision making and recommendation formulation in guidelines is unknown,^48^ but their potential effects must be kept in mind and made explicit to the extent possible.

While acknowledging the complex process of formulating recommendations in guidelines, EtD frameworks are a valuable tool and have led to vast improvements over “free-for-all” meetings where decision making criteria were selected in an *ad hoc* manner, and it was often unclear which considerations actually contributed to final decisions and their relative weight, what evidence was examined, and how and why the final recommendation or decision was arrived at.

### Applying clinical and public health frameworks to environmental health

Applying approaches for clinical medicine interventions to other scientific fields, including environmental health, is challenging. There are important differences both in the assessment of the quality (certainty) of the body of evidence, and in translating evidence to recommendations or decisions. Woodruff and colleagues^15^ note that the GRADE system and other evidence-based medicine approaches have limitations in terms of applicability to questions and decision making in environmental health. The reasons include: i) the need to combine human, animal and (sometimes) mechanistic evidence; ii) the paucity of randomized controlled trials (RCTs) and other types of experimental studies in humans due to ethical considerations; and iii) differences in the decision-making context (e.g., weighing the benefits and harms is different for clinical interventions than for unintended exposures to substances in the environment).^15^ Nonetheless, the constructs of the GRADE and WHO-INTEGRATE frameworks likely apply to environmental health interventions.

### Limitations of this analysis

The approach taken for this review and analysis has a number of limitations. The methods for identifying existing frameworks were limited: the systematic review included only reviews and did not include a search for all published EtD frameworks, and focused only on English language literature accessible through PubMed. Furthermore, many EtD frameworks are not published in the peer reviewed literature: rather, they are found on organizational web-sites and/or in organizational procedure manuals. For the review of key organizations, only a convenience sample was examined and EtD frameworks with important additional considerations or novel approaches may have been overlooked. However, it is unlikely that this approach missed key or significantly different frameworks in view of the dominance of GRADE and our consultation with experts on EtD frameworks.

Our analysis also has limitations related to the nature of the available data. Published information on the methods for developing frameworks was generally sparse, except for GRADE and WHO-INTEGRATE. The descriptions of the EtD criteria and how to operationalize them were also sparse for many frameworks. For some frameworks it was difficult to identify exactly which criteria were routinely included in the decision making process. While the publication might present a list or table of criteria, additional criteria might be mentioned in the adjacent text. However, the degree to which the guideline group or other decision maker addressed these additional criteria was often unclear.

## Conclusions and next steps

EtD frameworks are an extremely useful tool for recommendation formulation and decision making in healthcare and other scientific fields. They enable decisions to be made based on research evidence and with explicit consideration of a range of constructs, and they facilitate clear articulation of the rationale for decisions.

The principles underpinning evidence-informed, transparent and impactful decision making are the same across scientific fields, including environmental health. The GRADE EtD framework, along with WHO-INTEGRATE with its focus on contextual issues and inter-relationships, provide a useful starting point for consideration for decision-making for interventions in environmental health. Significant modifications will be needed, however, given the nature of the evidence base and the complex context in which environmental health interventions are designed, implemented, regulated and evaluated. The process for developing an EtD framework for environmental health interventions requires a broad range of experts, including not only environmental health scientists, but evidence synthesis and guideline methodologists, public health generalists, human rights and ethics experts, social scientists, and economists, among others. An iterative development process will be needed: a draft framework should be pilot-tested, revised and evaluated. The framework’s utility and impact on decision making, and the quality and impact of the resultant recommendations need careful evaluation, with the results used to develop future iterations.

## Data Availability

All data are available from the corresponding author.

## Annexes (online)

### Annex 1. Search strategy and inclusion criteria

((“Policy Making”[Mesh]) OR (“Decision Making”[Mesh])) AND ( ((framework[Title/Abstract]) OR (frameworks[Title/Abstract])) OR ((template[Title/Abstract]) OR (templates[Title/Abstract]) OR (tool[Title/Abstract]) OR (tools[Title/Abstract])))

Filters: English language, systematic review, last 10 years

The search was executed 7 January 2021 in the PubMed search engine for Medline.

### Annex 2. PRISMA flow diagram

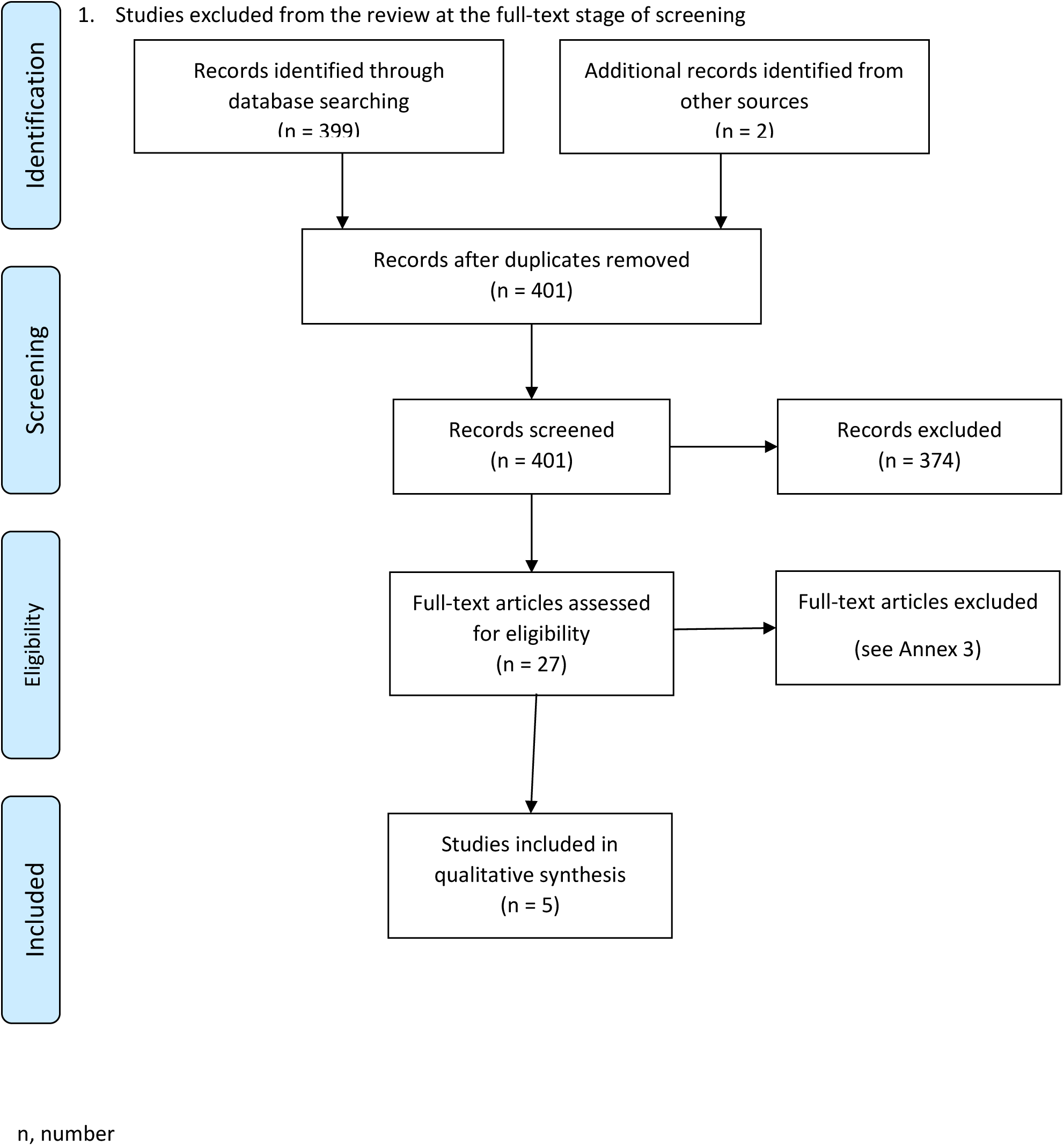

### Annex 3. Results of the search: full-text review

The search of PubMed performed on 7 Jan 2021. 399 citations were identified, 25 of which were reviewed in full text: these are listed below with the reason for exclusion.

Abbreviations: E, excluded citation; E2D, evidence-to-decision; I, included citation; LMICs, low- and middle-income countries; NA (Not applicable because the publication fulfilled inclusion criteria)

| Citation | Title | Include/exclude | Rationale for exclusion |
| --- | --- | --- | --- |
| Baltussen 2019 (1) | Multicriteria decision analysis to support health technology assessment agencies: Benefits, limitations, and the way forward | I | NA |
| Boivin 2018 (2) | Patient and public engagement in research and health system decision making: A systematic review of evaluation tools | E | Review of tools for evaluating patient and public engagement, not for decision making |
| Burchett 2012 (3) | National decision-making on adopting new vaccines: a systematic review | I | NA |
| Carroll 2013 (4) | Best fit framework synthesis: refining the method | E | Describes "best fit" method for qualitative synthesis; no EtD frameworks |
| Carvalho 2018 (5) | Capturing budget impact considerations within economic evaluations: A systematic review of economic evaluations of rotavirus vaccine in low- and middle-income countries and a proposed assessment framework | E | Checklist for budget impact analyses |
| Clark 2014 (6) | Discrete choice experiments in health economics: A review of the literature | E | Review of discrete chose experiments; no examination of EtD frameworks |
| Clark 2018 (7) | Measuring trade-offs in nephrology: A systematic review of discrete choice experiments and conjoint analysis studies | E | Review of discrete choice experiments in nephrology |
| Clarke 2016 (8) | The application of theories of the policy process to obesity prevention: a systematic review and meta-synthesis | E | Review of studies where theory underpins the policy development process or adoption of policies. Focus is on explaining the policies and not on what factors should be considered in decision-making. |
| Clifford 2017 (9) | What information is used in treatment decision aids? A systematic review of the types of evidence populating health decision aids | E | Focus on patient decision aids |
| Dodd 2019 (10) | Investigating the process of evidence-informed health policymaking in Bangladesh: a systematic review | E | Reviews factors linked to policy uptake or prioritization and how and why evidence is used; presents explanatory and descriptive information and not frameworks for normative decisions |
| Gentil 2015 (11) | A systematic review of socio-economic assessments in support of coastal zone management (1992-2011) | E | Describes concepts used for decision-making; presents descriptive information and not normative frameworks |
| Kolasa 2015 (12) | Pricing and reimbursement frameworks in Central Eastern Europe: a decision tool to support choices | E | Review revealed 33 pricing and reimbursement schemes; led to framework with two economic questions. Exclude as exclusively economic focus. |
| Kristensen 2019 (13) | Identifying the need for good practices in health technology assessment: Summary of the ISPOR HTA Council Working Group Report on Good Practices in HTA | E | Neither this paper or the full report (https://www.ispor.org/member-groups/councils-roundtables/health-technology-assessment-council) provides a review of EtD frameworks (referred to as "contextualization"). |
| Lam 2018 (14) | Decision support tools for regenerative medicine: Systematic review | E | Examines decision support tools for product development and manufacturing of cell and gene therapies in regenerative medicine; does not focus on clinical medicine, public health or environmental health |
| LaRocca 2012 (15) | The effectiveness of knowledge translation strategies used in public health: A systematic review | E | Review of evaluations of knowledge translation strategies and not the strategies themselves |
| Laurans 2013 (16) | Use of ecosystem services economic valuation for decision making: Questioning a literature blindspot | E | Focuses on the types and uses of Ecosystem Services economic Valuation (ESV), not on the criteria for decision making |
| Liverani 2013 (17) | Political and institutional influences on the use of evidence in public health policy. A systematic review | E | Examines factors that impact the use of evidence in policy-making |
| Mayer 2017 (18) | Costing evidence for health care decision making in Austria: A systematic review | E | Examines economic analyses and costing methods used in Austria; does not present EtD frameworks |
| Mustafa 2017 (19) | Decision making about healthcare-related tests and diagnostic test strategies. Paper 3: a systematic review shows limitations in most tools designed to assess quality and develop recommendations | I | NA |
| Sullivan 2015 (20) | What guidance are economists given on how to present economic evaluations for policymakers? A systematic review | E | Review of reporting guidance for economic analyses; does not deal directly with EtD concepts |
| Trapero-Bertram 2019 (21) | What attributes should be included in a discrete choice experiment related to health technologies? A systematic literature review | E | Review of attributes for discrete choice experiments; not a review of EtD frameworks |
| Tsoi 2013 (22) | Harmonization of reimbursement and regulatory approval processes: a systematic review of international experiences | E | Review of approaches to harmonize reimbursement and regulatory approval processes |
| Tsoi 2015 (23) | Systematic narrative review of decision frameworks to select the appropriate modelling approaches for health economic evaluations | E | Examines frameworks for selecting economic modelling approaches |
| Votruba 2018 (24) | A systematic review of frameworks for the interrelationships of mental health evidence and policy in low- and middle-income countries | E | Review of theories and frameworks explaining use of evidence to formulate policies in mental health in LMICs; does not review frameworks for decision-making |
| Wickremasinghe 2016 (25) | District decision-making for health in low-income settings: A systematic literature review | I | NA |
| **From other sources/ad hoc** |  |  |  |
| Domrumel 2020 (26) | Organizational aspect in healthcare decision-making: a literature review | E | Systematic review of individual criteria for decision-making (not a review of frameworks or sets of criteria) |
| Morgan 2018 (27) | Decision-making frameworks and considerations for informing coverage decisions for healthcare interventions: a critical interpretive synthesis. | I | NA |

